# Substantia nigra ferric overload and neuromelanin loss in Parkinson’s disease measured with 7T MRI

**DOI:** 10.1101/2021.04.13.21255416

**Authors:** Catarina Rua, Claire O’Callaghan, Rong Ye, Frank H. Hezemans, Luca Passamonti, P Simon Jones, Guy B Williams, Christopher T Rodgers, James B Rowe

**Affiliations:** Wolfson Brain Imaging Centre, Department of Clinical Neurosciences, University of Cambridge, Cambridge, UK; Department of Clinical Neurosciences, University of Cambridge, Cambridge, UK; Department of Psychiatry, University of Cambridge, UK; Brain and Mind Centre and School of Medical Sciences, Faculty of Medicine and Health, University of Sydney, Australia; Istituto di Bioimmagini e Fisiologia Molecolare (IBFM), Consiglio Nazionale delle Ricerche (CNR), Milano, Italia; MRC Cognition and Brain Sciences Unit, University of Cambridge, Cambridge, UK

**Keywords:** 7T MRI, Parkinson’s disease, Substantia Nigra, Iron, Neuromelanin

## Abstract

**Background:** Vulnerability of the substantia nigra dopaminergic neurons in Parkinson’s disease is associated with ferric overload, leading to neurodegeneration with cognitive and motor decline. Here, we quantify iron and neuromelanin-related markers *in vivo* using ultra-high field 7-Tesla MRI, and examine the clinical correlates of these imaging assessments.

**Methods:** Twenty-five people with mild-to-moderate Parkinson’s disease and twenty-six healthy controls underwent high-resolution imaging at 7-Tesla with a T_2_*-weighted sequence (measuring susceptibility-χ and R_2_*, sensitive to iron) and a magnetization transfer-weighted sequence (MT-w, sensitive to neuromelanin). From an independent control group (N=29), we created study-specific regions-of-interest for five neuromelanin- and/or iron-rich subregions within the substantia nigra. Mean R_2_*, susceptibility-χ and their ratio, as well as the MT-w contrast-to-noise ratio (MT-CNR) were extracted from these regions and compared between groups. We then tested the relationships between these imaging metrics and clinical severity.

**Results:** People with Parkinson’s disease showed a significant ~50% reduction in MT-CNR compared to healthy controls. They also showed a 1.2-fold increase in ferric iron loading (elevation of the 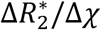 ratio from 0.19±0.058ms/ppm to 0.22±0.059ms/ppm) in an area of the substantia nigra identified as having both high neuromelanin and susceptibility MRI signal in healthy controls. In this region, the ferric-to-ferrous iron loading was associated with disease duration (β=0.0072, p_FDR_=0.048) and cognitive impairment (β=−0.0115, p_FDR_=0.048).

**Conclusions:** T_2_*-weighted and MT-weighted high-resolution 7T imaging markers identified neurochemical consequences of Parkinson’s disease, in overlapping but not-identical regions. These changes correlated with non-motor symptoms.

## 1. Introduction

Iron accumulation and loss of pigmented neuromelanin cells in the substantia nigra are key parts of the complex neuropathology of Parkinson’s disease (1–6). While iron is essential for normal brain homeostasis, it can become toxic by the generation of free radicals promoted by redox reactions, converting ferrous Fe^2+^ to the ferric Fe^3+^ form (7). In Parkinson’s disease, iron overload and increased oxygen free radical formation induce lipid peroxidation of cell membranes (8), promoting dopaminergic neurodegeneration (9). In contrast, neuromelanin can provide a neuroprotective pathway by chelating iron and hence preventing damage by Fe^3+^ (10,11). However, in Parkinson’s disease, the buffering capacity of neuromelanin may be exceeded, leading to neurotoxicity, oxidative stress, neuroinflammation, and ultimately loss of neurons (10,12).

These pathological features in the substantia nigra of patients with Parkinson’s disease have been extensively confirmed in *post mortem* studies (1,2,4,6,13). However, characterising these changes and distinguishing different types of iron *in vivo* remains a challenge. Recently, specialised MRI sequences have been developed to characterise tissue iron and changes in neuromelanin content. With sufficient resolution and sensitivity, these could potentially be used to identify functionally distinct sub-regions of the substantia nigra. The ventral portion of the substantia nigra, known as the substantia nigra *pars reticulata* (SNr), has high concentration of iron in healthy subjects. The dorsal portion, known as the substantia nigra *pars compacta* (SNpc), is rich in neuromelanin, the pigmented by-product of dopamine and noradrenaline metabolism (11,14).

The dopaminergic-rich SNpc can be imaged *in vivo* using a neuromelanin-sensitive “magnetisation-transfer weighted” (MT-w) MRI sequence (15–17). The MT-w signal intensity correlates with the location and density of the neuromelanin content in older adults (18). Alternative sequences have been used to observe changes in iron in the healthy brain and in disease (19,20) by measuring the T_2_* transverse relaxation time (or its inverse rate, R_2_*) or the local tissue magnetic susceptibly χ using quantitative susceptibility mapping (QSM) (21–23). Susceptibility weighted imaging (SWI) uses post-processing of T_2_*-w images, with the magnitude and filtered phase information providing image contrast from iron storage (24). SWI has been shown to provide unique contrast to identify the appearance of the nigrossome-1 (N1-sign) in healthy controls, and its loss in patients with Parkinson’s disease (24).

Susceptibility imaging is sensitive to any molecule that generates an MR phase shift. In contrast to SWI, both QSM and R_2_* provide quantitative measures of susceptibility effects. However, they are unable on their own to differentiate the sources of susceptibility, and therefore an adjunctive method is required. One approach is to capitalise on the fact that ferrous and ferric irons show differences in relaxivity per unit concentration (25), which can be estimated from the R_2_*-to-χ ratio. We therefore take the R_2_*-to-χ ratio as a proxy of ferric vs. ferrous loading, and use this ratio as a biomarker for iron accumulation in Parkinson’s disease.

In this study, we used high resolution dedicated MRI sequences for *in vivo* imaging of neuromelanin with high resolution (0.08 mm^3^) and iron (0.34 mm^3^) using ultra-high field 7T, based on magnetisation transfer (26) and multi-echo T_2_*, respectively, with nigral regions of interest derived from an independent cohort of age-matched healthy controls (27). The contrast-to-noise ratio (CNR) of the MT signal was interpreted as a measure of neuromelanin content, whereas χ and R_2_* were used to quantify iron in the tissue. We investigated (1) the molecular changes of neuromelanin and iron in sub-regions of the substantia nigra in Parkinson’s disease compared to healthy controls, and (2) the association between imaging markers and cognitive and motor functions in the patient group. We hypothesised that people with Parkinson’s disease would show decreased MT signal contrast and increased χ and R_2_* in the SNpc. Since ferric iron demonstrates higher susceptibility and R_2_* values than ferrous iron at the same concentration in tissue (25,28) we also expected an increase of the R_2_*-to-χ ratio in the SNpc.

## 2. Methods

### 2.1 Participants

Twenty-six healthy controls (HC, 15 male, age 65±5 years) and twenty-five participants with Parkinson’s disease (PD, 18 male, age 67±7 years) were recruited via the University of Cambridge Parkinson’s disease research clinic and the Parkinson’s UK volunteer network. The study was approved by the Cambridge Research Ethics Committee (16/EE/0084; 10/H0308/34) and all participants provided written informed consent in accordance with the Declaration of Helsinki. Patients met the United Kingdom Parkinson’s Disease Society Brain Bank criteria and did not have dementia, based on clinical impression and mini-mental state examination score (MMSE) >26/30 (29) (Table 1). They were Hoehn and Yahr stage 1.5-3, with no contraindications to 7T MRI. None had current impulse control disorders, based on clinical impression and the Questionnaire for Impulsive-Compulsive Disorders in Parkinson’s Disease (QUIP-Current Short) screening tool (30). Levodopa equivalent daily dose (LEDD) scores were calculated (31) and patients were scanned on their regular medications. Control participants were matched for age, sex and education, and were screened for a history of other neurological or psychiatric disorders, current psychoactive medications, and contraindications to 7T MRI. Two control participants were excluded due to abnormal incidental findings on their T_1_-w structural images.

**Table 1:**
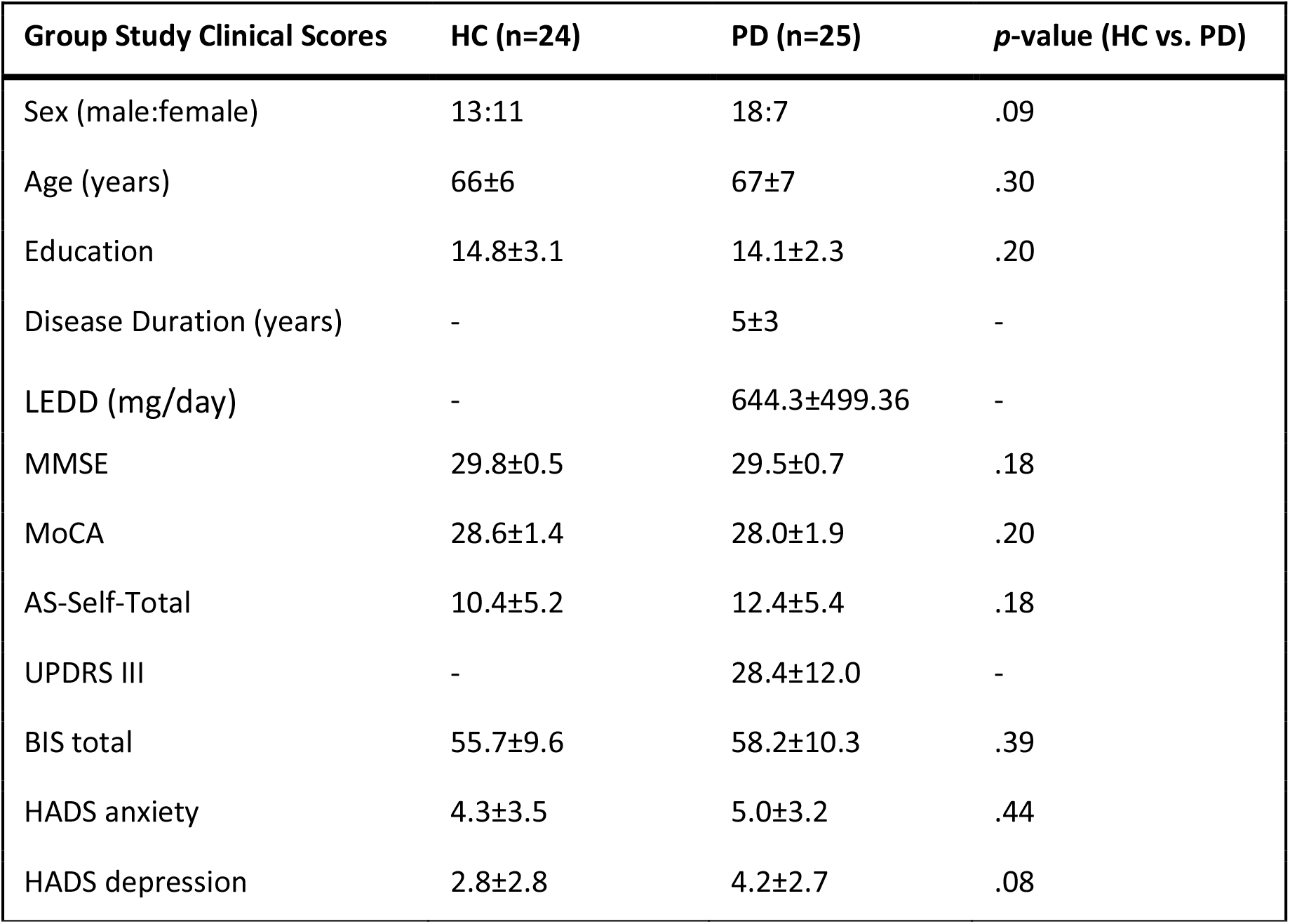
Average ± standard-deviation values for the demographics and clinical assessments of the healthy controls (HC) and patients with Parkinson’s disease (PD). Group differences in sex were tested using the chi-squared test. For all other group comparisons, a two-tailed two-sample t-test was used. P-values uncorrected for multiple comparisons. *Abbreviations: AS - Apathy Scale; BIS - Barratt Impulsiveness Scale; HADS - Hospital Anxiety and Depression scale; MMSE - mini-mental state examination score; MoCA - Montreal cognitive assessment; UPDRS - Unified Parkinson’s Disease Rating Scale; LEDD - Levodopa equivalent daily dose*.

| Group Study Clinical Scores | HC (n=24) | PD (n=25) | p-value (HC vs. PD) |
| --- | --- | --- | --- |
| Sex (male:female) | 13:11 | 18:7 | .09 |
| Age (years) | 66±6 | 67±7 | .30 |
| Education | 14.8±3.1 | 14.1±2.3 | .20 |
| Disease Duration (years) | - | 5±3 | - |
| LEDD (mg/day) | - | 644.3±499.36 | - |
| MMSE | 29.8±0.5 | 29.5±0.7 | .18 |
| MoCA | 28.6±1.4 | 28.0±1.9 | .20 |
| AS-Self-Total | 10.4±5.2 | 12.4±5.4 | .18 |
| UPDRS III | - | 28.4±12.0 | - |
| BIS total | 55.7±9.6 | 58.2±10.3 | .39 |
| HADS anxiety | 4.3±3.5 | 5.0±3.2 | .44 |
| HADS depression | 2.8±2.8 | 4.2±2.7 | .08 |

Patients were assessed using the motor component of the Movement Disorder Society Unified Parkinson’s Disease Rating Scale (MDS-UPDRS-III; (32)), and both patients and healthy controls underwent cognitive screening with the mini-mental state examination (MMSE; (33)), Montreal cognitive assessment (MoCA; (34)). Self-rated questionnaires were used to assess anxiety and depression (Hospital Anxiety and Depression Scale; HADS (35)), impulsivity (Barratt Impulsiveness Scale; BIS-11 (36)) and apathy (Apathy Scale; (37)).

To create a template and select regions-of-interest, an independent sample of 30 additional age-, sex- and education-matched healthy controls (HCi, 17 male, age 67±8 years) was collected as part of the same study protocol. One participant was excluded because of excessive movement during scanning, resulting in 29 healthy controls in the HCi group.

### 2.2. MRI acquisition

MR imaging was acquired on a 7T Magnetom Terra scanner (Siemens, Erlangen, Germany) equipped with a 32-channel receive and circularly polarized single-channel transmit head coil (Nova Medical, Wilmington, USA). For iron imaging, 3D multi-echo, 0.7mm isotropic T_2_*-weighted images were acquired transversely for QSM, R_2_* mapping and SWI: TE1/TR=4.68/27ms; 6 echoes, echo-spacing=3.24ms, FA=15º; BW=430Hz/px; acceleration-factor=2×2; FOV=224×196×157mm^3^; ~7min TA; and Roemer coil combination (38). An MT-w turbo flash sequence was used for neuromelanin imaging as described in (26,27). Briefly, a train of 20 RF-pulses at 6.72ppm off-resonance preceded a Turbo-FLASH readout: TE/TR=3.44/1251ms, flip-angle=8º, voxel-size=0.4×0.4×0.5mm^3^, BW=300Hz/px, no acceleration, slices=112, 14.3%-oversampling, ~7min TA. The MT-preparation pulses were calibrated to 420º FA at the middle of the pons. Two MT sequence repeats (MT-on) were acquired on the HC and PD groups and an additional MT-on repeat was acquired in the HCi group; a repeat without off-resonance pulses was also acquired in all three groups (MT-off). For anatomical co-registration, 0.7mm isotropic MP2RAGE images were acquired as described in (39).

### 2.3. Data pre-processing

In-house MATLAB scripts (R2018b, The MathWorks, Massachusetts, United States) together with ANTs v2.2.0 and FSL v5.0.10 were used for image pre-processing (processing scripts available upon request). QSM maps were estimated from the T_2_*-w scans using the multi-scale dipole inversion algorithm in QSMbox (40) with the pipeline described in (23) but with the regularization parameter set to 10^3.3^. R_2_* was estimated with the ARLO algorithm (41). To detect the relaxivity effects of ferrous (Fe^2+^) and ferric (Fe^3+^) components (25), the 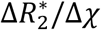 ratio was measured in each voxel *i*, 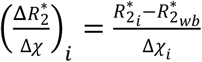, by the ratio of the normalised R_2_^*^ maps (referenced to the whole brain R_2_* value 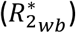) and the Δ*χ* measured by QSM (MSDI QSM-processing references χ to the whole brain χ value).

To calculate SWI maps, for each echo the raw phase was filtered with a homodyne high-pass filter (filter strength: 6.2%). All echoes were then temporally fitted, and a normalized phase mask, *φ*_*M*_, was generated according to equation 2 of (42) under the assumption that the phase of interest is negative. SWI maps were computed by multiplying the T_2_*-map with the fourth power of the normalized phase mask 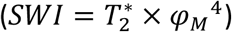.

All MT scans (MT-on and MT-off) were pre-processed as previously described (27). Briefly, data was first bias field corrected. Then the MT-on images (2 repeats for HC and 3 repeats for HCi groups) were combined with the customised antsMultivariateTemplateContruction2 function setting the Laplacian sharpening off in the averaging step.

The T_1_-w MP2RAGE image was generated offline from the raw magnitude and phase of the two inversion times (43). Then, were bias-field corrected with ANTs and segmented with SPM12 (v7219) and skull-stripped.

### 2.4. Analysis of independent control dataset

A T_1_-w driven, cross-modality co-registration pipeline was built to warp the MT and the QSM data from the HCi group into the isotropic 0.5mm ICBM152 T_1_-w asymmetric brain template (44) (Figure 1). On a within-subject level, the T_1_-w images were first independently registered to the combined MT-on image (rigid transform, via the MT-off image (27)), and to the first echo of the T_2_*-w scan (rigid transform using the cross-correlation metric with weight 1 and window radius 4 mm). A T_1_-w structural template was created from the individual T_1_-w data using a three step registration process: 1) rigid, 2) affine, and 3) hierarchical nonlinear diffeomorphic registration with greedy symmetric normalisation (SyN) algorithm (45) at 5 resolution levels. The study template was then registered to the ICBM 152 2009b standard image following an identical 3-step registration pipeline with 4 resolution levels. Details for each registration step are described in (27).

**Figure 1:**
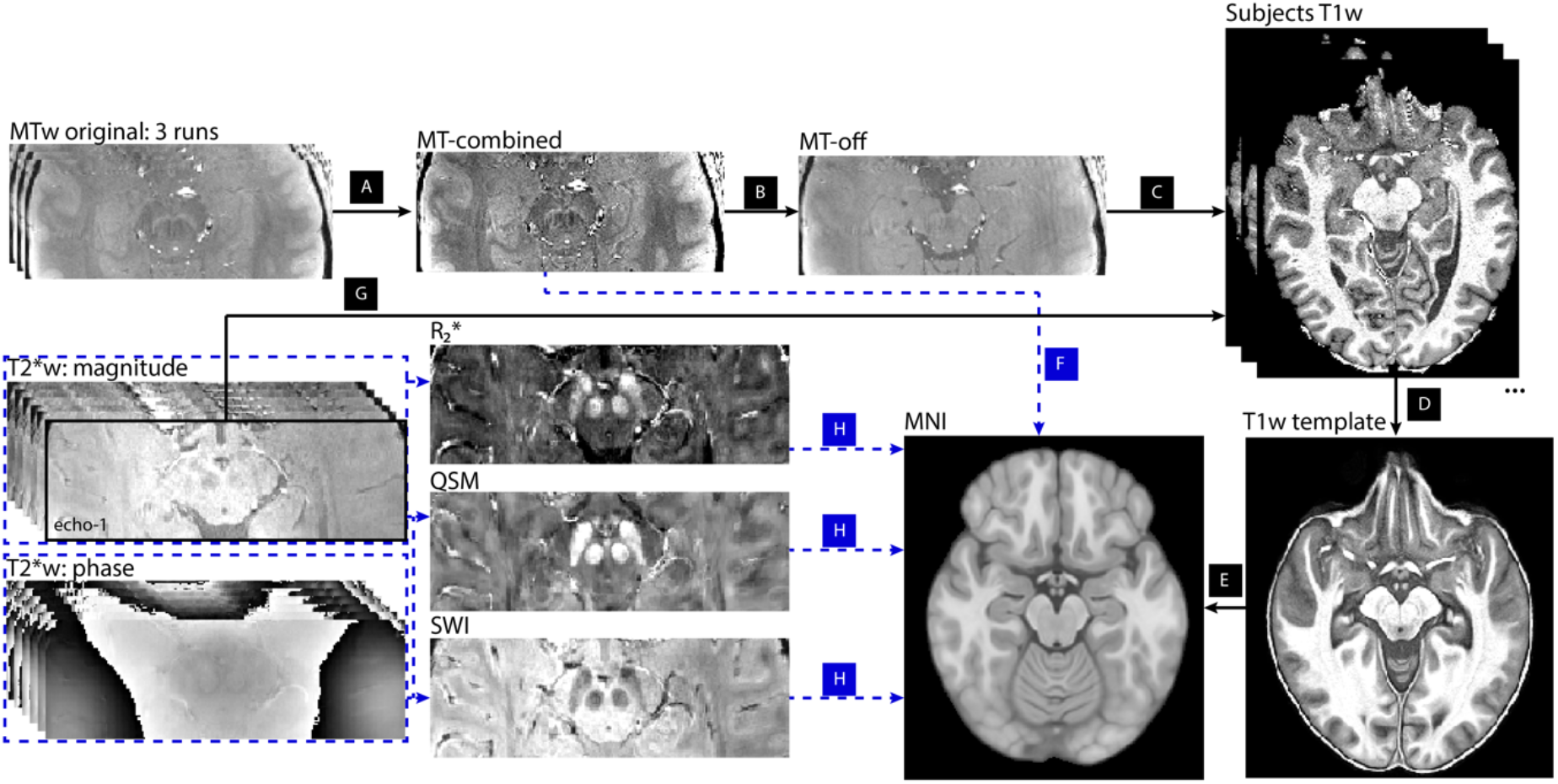
Registration pipeline for the MT-w and T_2_*-w data from subject space to the MNI ICBM152 standard brain. For the MT-w data, (A) the individual scans were first combined and registered to the MT-off scan. (B) The latter was used to find the registration between MT space and T_1_-w space (C). The skull-stripped T_1_-w scans from all subjects were used to create a study-wise template (D) which was then mapped to MNI space via a three-step non-linear registration (E). One single registration was applied on the MT-combined image which included the 4 linear and 2 non-linear transforms to bring the data to the standard space (F). The first echo of the T_2_*-w scans was used to register the T_2_*-w data into each subjects’ T_1_-w scan (G). The subsequent registration steps to standard space were identical to the MT-w data. The generated QSM, R_2_* and SWI maps were brought to MNI space by performing a single registration including one linear and two non-linear transforms (H).

All the participants’ combined MT-on images were registered in one step to ICBM 152 space with four linear transformation matrices and two non-linear deformations (Figure 1). Similarly, the QSM, R_2_* and SWI maps were registered in one step to ICBM 152 template via three linear and two non-linear transformations.

### 2.5. Generation of substantia nigra regions-of-interest

We applied a semi-automated threshold-based method to segment the substantia nigra independently in the MT-on and QSM images, as it has been validated for the substantia nigra and smaller brain regions such as the locus coeruleus (27,46,47). The N1-sign was segmented in SWI images with a similar approach.

Since MT-on, and QSM and SWI images show different contrast characteristics in the substantia nigra (48), two different search areas were defined. The search area for the substantia nigra was manually defined bilaterally in ITK-SNAP software v3.6.0 (49) within the midbrain portion of the brainstem ROI defined from MNI template brainstem substructure segmentation in FreeSurfer 6.0 (50) (Supplementary Information Figure 1). The slices that showed hyperintensities in the substantia nigra region were selected.

Cylindrical reference regions were defined in the left and right hemispheres (diameter 3.5 mm, 4 mm height) in the lateral crus cerebri adjacent to the substantia nigra, starting from the most caudal slice with visible hyperintensity in the anatomic region of the substantia nigra in the MT-on and QSM images (z=105 to z=112 of the standard ICBM 152 MNI space - Supplementary material Figure 1D).

Then, for each subject and image contrast separately, the substantia nigra voxels were thresholded at a level defined by the hemispheric specific reference region: *T* = *AV*_*REF*_ + *α* × *SD*_*REF*_, where *α* was set to 5 for the QSM and 4 for the MT-on images. For the SWI images, because the contrast within the ref region is similar N1-sign of the substantia nigra, *α* was set to −1.5. A resulting binary map for each subject and contrast was built based on the signal that survived the threshold *T*. Each participant’s map was added to create a probability density map of the MT-on, QSM and SWI based substantia nigra segmentation, and were thresholded (at 5% for MT-on, 35% for QSM, and 25% for SWI) to obtain the final bilateral ROIs (‘MTROI’, ‘QSMROI’ and ‘SWIROI’, respectively - Figure 2). The SWIROI required further manual adjustments to include only voxels within the MTROI or the QSMROI and not the crus cerebri (Supplementary Figure 3).

**Figure 2:**
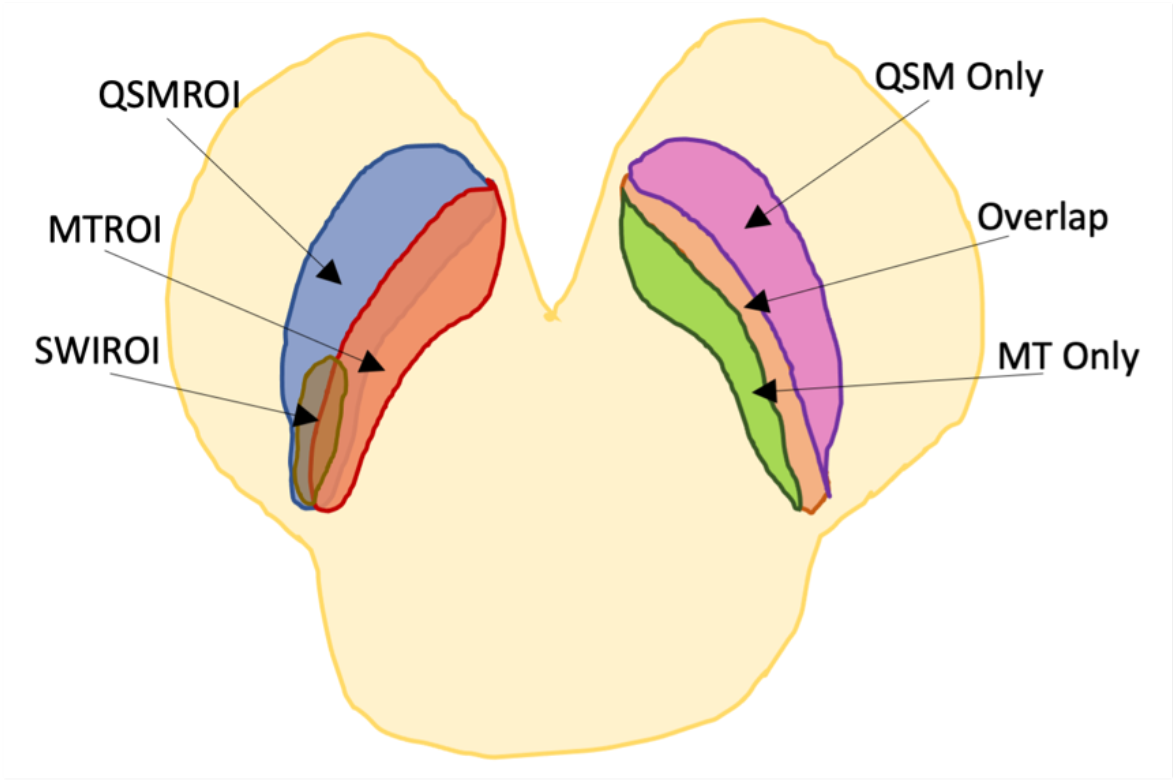
ROI definition in the substantia nigra used in this study identified from an independent cohort. Two ROIs were defined based on the substantia nigra QSM hyperintensity (‘QSMROI’ – blue) and MT-w hyperintensity (‘MTROI’ – red). The ‘SWIROI’ (brown) was defined as the hyperintensity in the SWI images within the MTROI and QSMROI. Then, three new ROIs were defined: the ‘Overlap’ ROI included voxels that were common to the QSMROI and MTROI (orange); the ‘QSMOnly’ ROI contained voxels that were part of the QSMROI and not of the MTROI (purple); the ‘MTOnly’ ROI included voxels that were exclusive to the MTROI and not to the QSMROI (green).

Three new bilateral sub-ROIs were defined (Figure 2): the ‘Overlap’ ROI included voxels that were part both of MTROI and QSMROI; the ‘MTOnly’ ROI included voxels of the MTROI which were not part of the Overlap ROI; the ‘QSMOnly’ ROI included voxels of the QSMROI which were not part of the Overlap ROI.

### 2.6. Signal extraction from the substantia nigra

In the HC and PD groups, the neuromelanin (combined MT-on) and iron (QSM and R_2_*) maps were mapped to ICBM152 space via a similar pipeline to the HCi described in section 2.4, with an unbiased T_1_-template created from the individual T_1_-w MP2RAGE scans from the participants both in the HC and PD groups.

In standard space, the MT-CNR was computed on the combined MT-on images within an ROI by extracting the difference between the mean signal of the hemisphere specific ROI 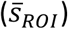 and the mean intensity in the reference region of the corresponding hemisphere 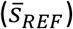, divided by the standard deviation of the reference signal 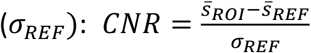. In addition, the average QSM-Δ*χ*, R_2_* and 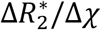 ratio were extracted from all the ROIs.

### 2.7. Statistics

Statistical tests were performed in R (v1.3.1093). For each region-of-interest (described in section 2.5), distributions of each imaging metric 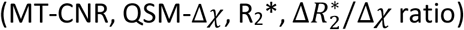 were evaluated by fitting data with normal, log-normal, gamma and logistic model distributions. The goodness-of-fit for the parametric distributions were calculated and the distribution which showed the lowest Akaike Information Criterion was then used on a general linear mixed effects model (glmer function in R). The 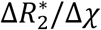 ratio was fitted with a gamma distribution whereas for all other metrics the Gaussian distribution resulted in the best fit.

To compare patients and control imaging metrics, the glmer model included group as a fixed effect of interest, hemisphere as a fixed effect of no interest, with a random effect of participant on the intercept. For statistical inference on the fixed effects, p-values were obtained by computing an analysis of deviance table on the fitted model (51). Receiver operating characteristic (ROC) analysis was performed and the area under the curve (AUC) was calculated to evaluate the overall diagnostic performance of each imaging metric in differentiating patient and control groups.

To test for associations with clinical features, linear models were fitted on the imaging data (averaged across both hemispheres) of the patients, separately for each clinical variable (disease duration, MoCA, UPDRS-III, AS-self-Total, BIS-Total, HADS-anxiety, HADS-depression), with age added as a nuisance covariate. To assess laterality effects on the patients, a clinical motor symptom laterality index (MAI) was calculated from the UPDRS-III scores and evaluated against imaging 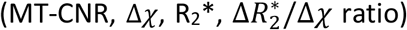 asymmetry indexes (details provided in supplementary information). Linear models were fitted on each of the imaging asymmetry indexes with MAI as a fixed effect and age as a covariate of no interest. False discovery rate (FDR) corrected p-values were extracted from the fits. The corrected p-value threshold for significance was 0.05.

## 3. Results

Demographic and clinical characteristics are shown in Table 1. The groups were matched for age, sex, education, and there were no significant difference across the cognitive screening measures or behavioural questionnaires. Parkinson’s disease duration was 5±3 years.

### 3.1. Neuromelanin and iron maps of healthy controls and Parkinson’s disease

Figure 3 shows axial and coronal views of the subject average MT, QSM, R_2_* and SWI maps in the substantia nigra region. The right Overlap ROI is outlined in green for all images. 53% of the MTROI voxels and 19% of the QSMROI voxels are included in the Overlap ROI. In controls, the location of the Overlap ROI is shown with respect to the N1-sign as observed in the SWI image (red outline - Figure 3D; approx. 25% and 31% shared voxels, respectively; Dice=0.3).

**Figure 3:**
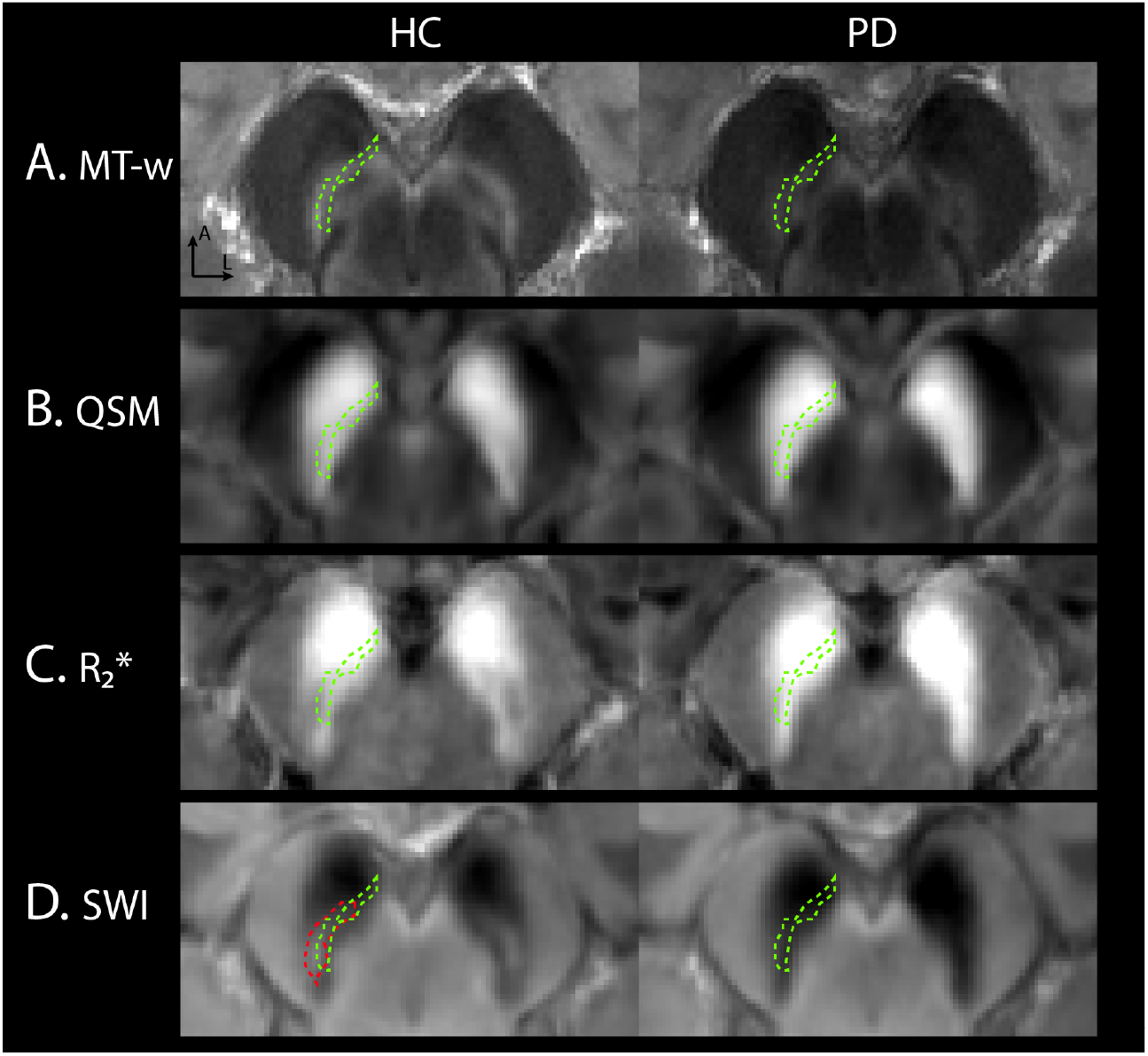
Axial slices of the group average maps showing the substantia nigra region for HC and PD groups for (A) MT-w, (B) QSM, (C) R_2_*, (D) SWI. Green outline shows the location of the same right Overlap ROI in all the images. In (D) the right nigrosome-1 (N1) sign is delineated in red.

MT images show an overall reduction of signal in the substantia nigra in participants with Parkinson’s disease compared to healthy controls. For iron-related measures, in participants with Parkinson’s disease, the N1 sign is not evident in the SWI images (Figure 3D), which is associated with a local increase in iron concentration as observed by the increased signal in QSM and R_2_* in the Overlap ROI (Figure 3B,C).

### 3.2. Group statistics

Compared to controls, participants with Parkinson’s disease had reduced neuromelanin signal in all nigral regions-of-interest as measured by MT-CNR (Figure 4A; and Supplementary Information Table 1). This reduction was largest in MT-based (MTROI and MTOnly) and Overlap regions (e.g. MT-CNR_HC_ = 4.6±2.1 and MT-CNRPD = 2.4±1.8 in the Overlap region) compared to QSM regions. Similarly, QSM-Δ*χ* increased 1.4-fold in MT-driven ROIs and in the Overlap ROI (e.g. Δ*χ*_HC_ = 0.12±0.03 ppm and Δ*χ*_PD_ = 0.15±0.03 ppm in the Overlap ROI, p < 0.0001) but did not reach statistical significance in the QSMROI, QSMOnly or SWIROI regions (Figure 4B). R_2_* showed similar statistics to QSM (Supplementary Information Table 2). The boxplots for the 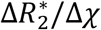 ratio show a local significant increase (p = 0.0048) within the Overlap ROI (Figure 5): compared to controls, the 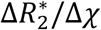 ratio was, on average, 0.031 ms/ppm higher in the patient group. Similar significant hemispheric differences were found in patient and control data in all imaging metrics (e.g. SWIROI of Figure 5, p<0.0001).

**Figure 4:**
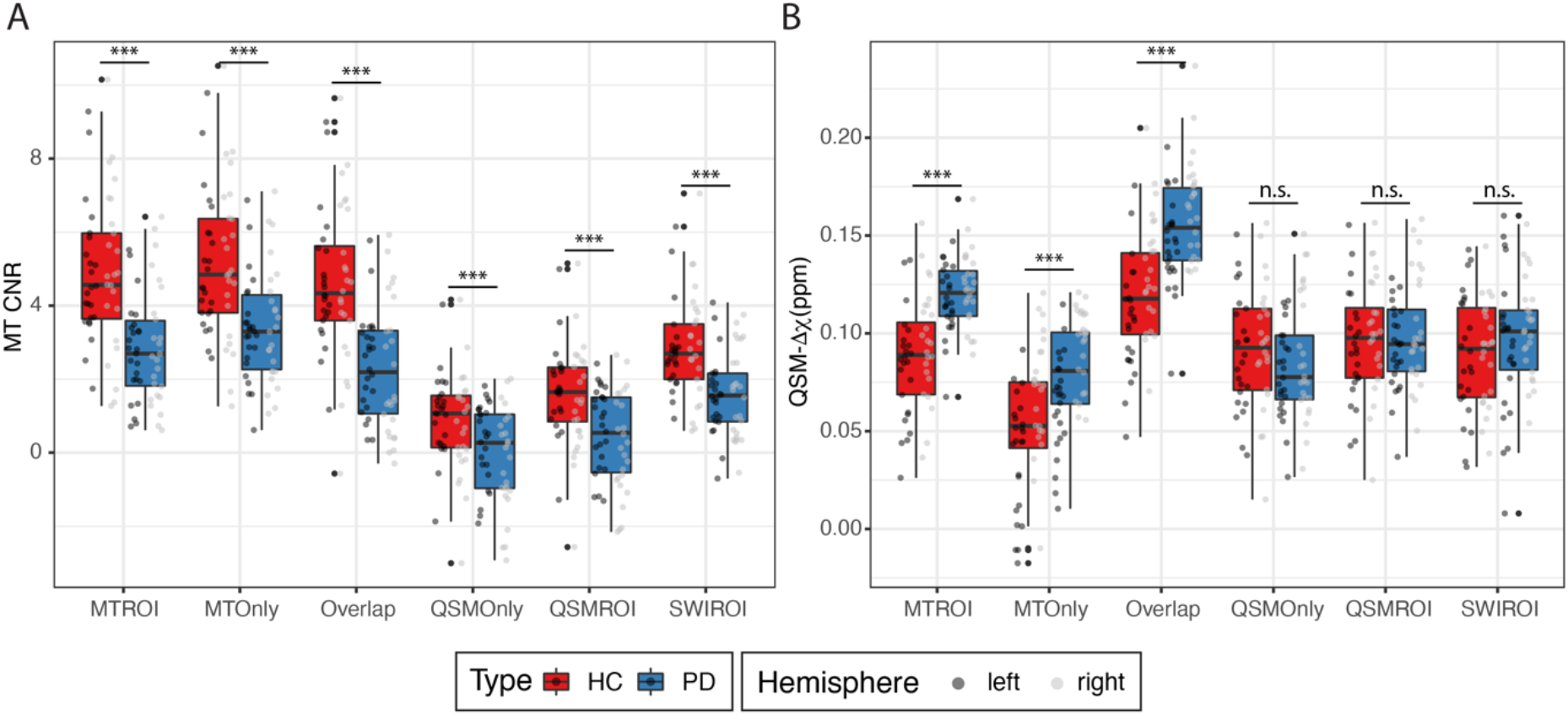
Boxplots of the MT-CNR (A) and QSM-Δ*χ* (B) in healthy controls (HC, red) and patients with Parkinson’s disease (PD, blue) in the six sub-regions the substantia nigra. Individual subject measures from the left and right hemispheres are overlaid as scatter points. Statistical significance: *** p<0.0001; ** p<0.01; n.s. p>0.05.

**Figure 5:**
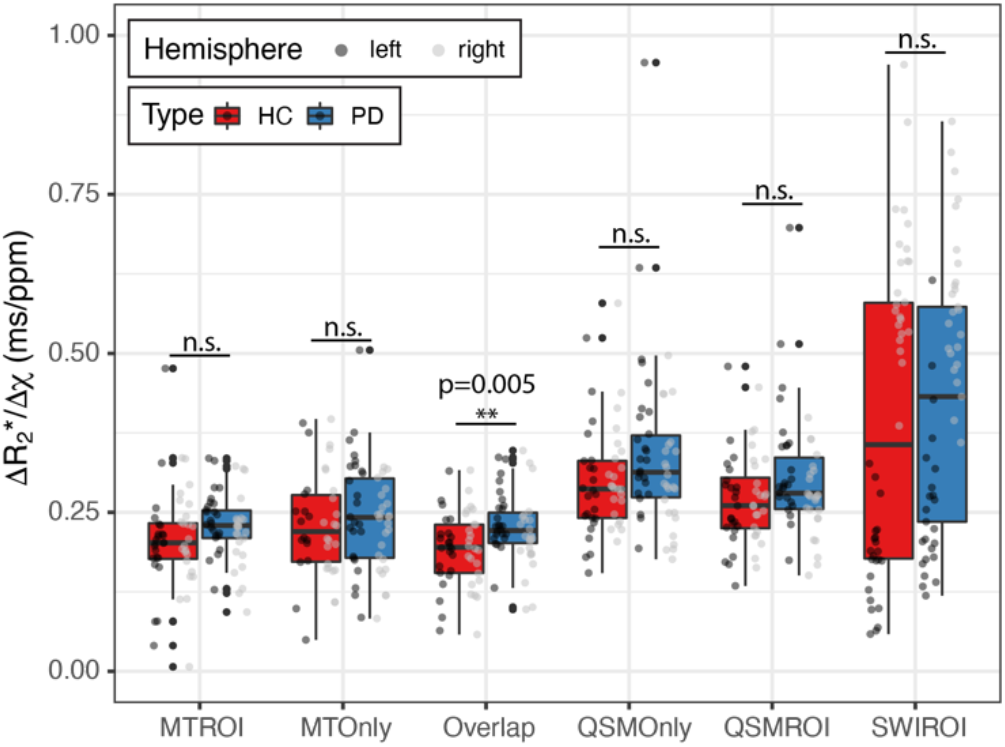
Boxplots of the 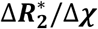 ratio in healthy controls (HC, red) and patients with Parkinson’s disease (PD, blue) in the six sub-regions the substantia nigra. Individual subject measures from the left and right hemispheres are overlaid as scatter points. Statistical significance: *** p<0.0001; ** p<0.01; n.s. p>0.05.

### 3.3. ROC analysis

Receiver operating characteristic curves for the MT-CNR, QSM-Δ*χ* and 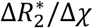 ratio are illustrated in Figure 6. Compared to the other regions-of-interest, the AUC values for the Overlap ROI were the highest when using the MT-CNR data (AUC=0.81; p<0.0001) and the QSM-Δ*χ* data (AUC=0.83; p<0.0001) in differentiating patients with Parkinson’s disease from healthy subjects. This indicates that, in the nigra the MT and QSM data share complementary information for differentiating patients from controls. For the 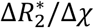 ratio the Overlap ROI also showed the largest AUC values (AUC=0.67; p=0.002). The SWIROI defining the N1-sign showed lower AUC values compared to the Overlap ROI in the three imaging metrics (Figure 6).

**Figure 6:**
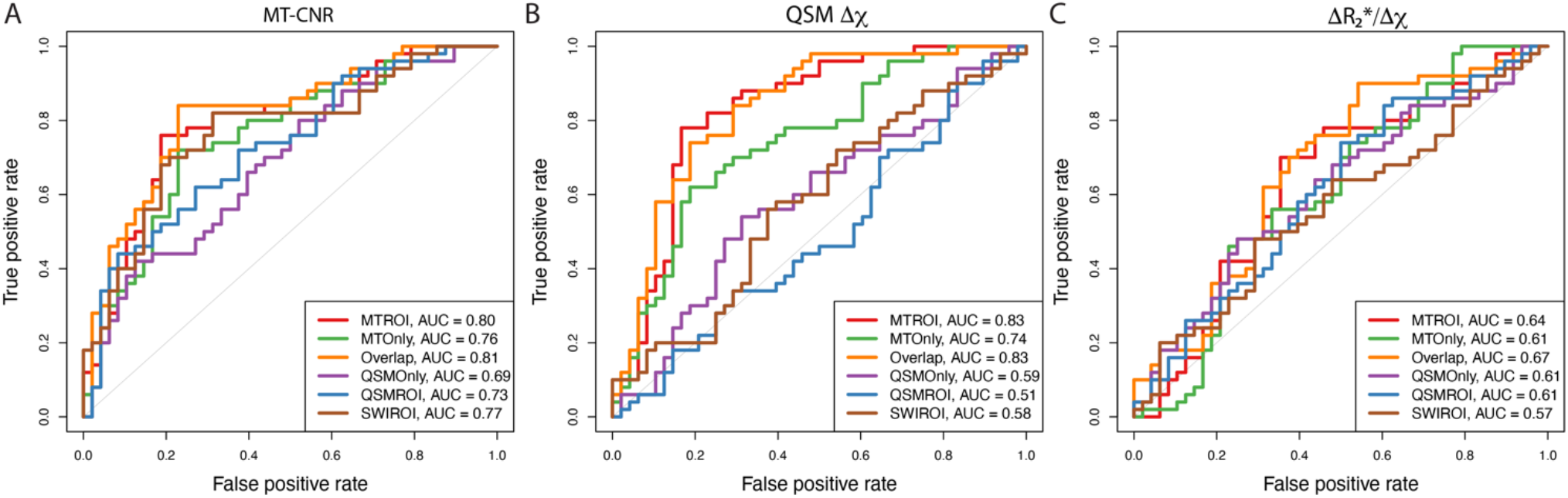
ROC plots for (A) MT-CNR, (B) QSM-Δ*χ* and (C) 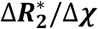 ratio discriminating patients with Parkinson’s disease and healthy controls based on data extracted from the six sub-regions of the substantia nigra. Data from both hemispheres included in the analysis.

### 3.4. Associations between imaging and clinical measures

Disease duration showed a significant positive association with the 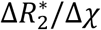 ratio in all ROIs including the overlap ROI (β=0.0072; p_fdr_ = 0.048) (Figure 7A), but not the MTOnly ROI (Supplementary Information Figure 4). Neither QSM nor R_2_* alone showed statistically significant associations with disease duration.

**Figure 7:**
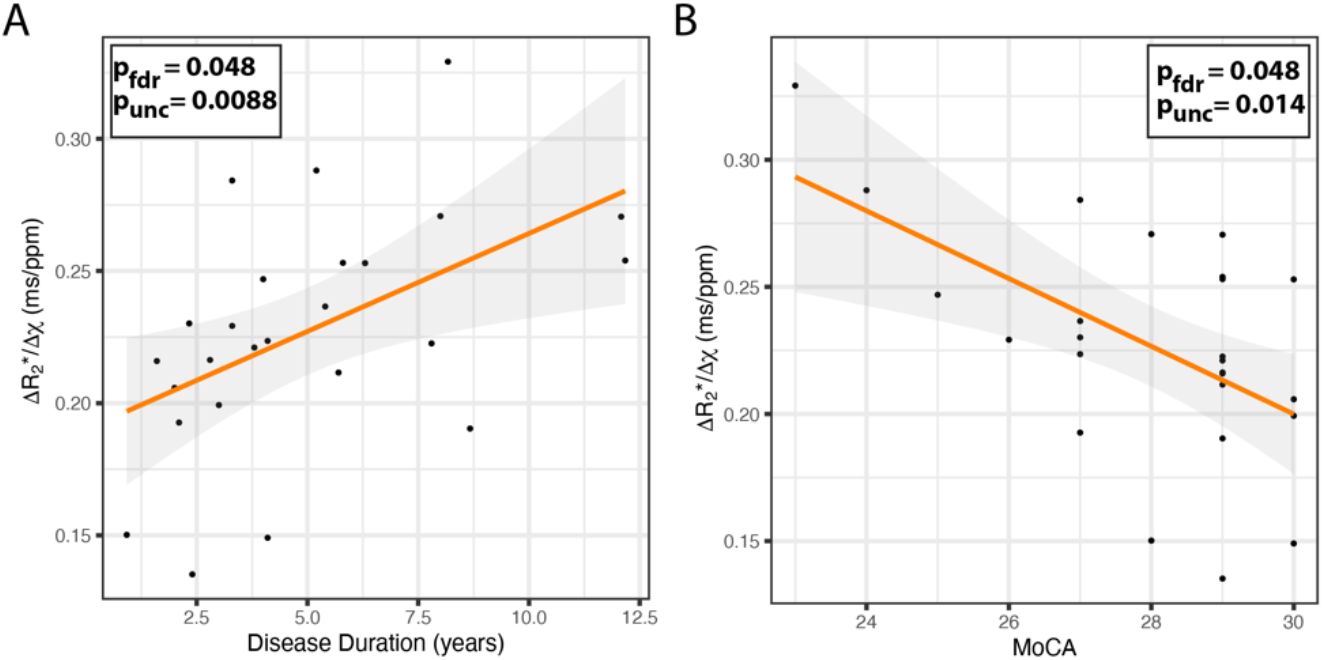
Relationship between the 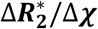 ratio and clinical characteristics of disease duration in years (A) and MoCA score (B) in the Parkinson’s disease group in the Overlap ROI. Uncorrected and FDR-corrected p-values are reported in each plot for the linear fits.

Increased 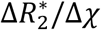 ratio was associated with lower MoCA scores in patients within the neuromelanin-rich substantia nigra (Supplementary Information Figure 5), reaching significance in the Overlap ROI (Figure 7B): β=-0.011; p_fdr_ = 0.048. Imaging measures showed no statistically significant associations with motor severity (UPDRS-III) after correction for multiple comparisons (p_fdr_ >0.3), apathy scale (p_fdr_ >0.3), Barrat impulsiveness scales (p_fdr_ >0.2), and anxiety and depression scales (p_fdr_ >0.3). No significant associations were found when evaluating the imaging laterality indexes against clinical motor symptom laterality indexes in all subregions of the substantia nigra.

## 4. Discussion

This study reveals the impact of Parkinson’s disease on neuromelanin (by MT contrast) and iron (by susceptibility quantities - QSM and R_2_*), using ultra-high field high-resolution magnetic resonance imaging of the substantia nigra. We defined two main regions-of-interest. In healthy individuals, MT-CNR was highest in the inferoventral region of the substantia nigra, whereas increased susceptibility was greatest in the superolateral region. As reported in previous imaging studies (48,52,53), these two regions represent to some degree the difference between the classically defined *pars compacta* and *pars reticulata*.

The *pars compacta* has closely packed pigmented neurons, while the *pars reticulata* shows scattered non-pigmented neurons in a reticular neuropil (54). Our 7T high-resolution imaging showed the additional definition of subregions associated to the substantia nigra based on the MT and QSM measures. We found that a sub-region showing increase of both these contrasts, the Overlap ROI, in healthy controls consistent with the anatomical location of the ventral portion of the SNpc (vSNpc), was the most sensitive to differentiating Parkinson’s disease as supported by the highest AUC on ROC analysis (Figure 6). This demonstrates that proxy MRI measures of neuromelanin loss and increased iron accumulation are complementary indices that help discriminating patients with Parkinson’s disease from controls. A similar three layer organisation has been proposed by Cosottini et al. (55).

Brammerloh et al. (56) demonstrated the dominant R_2_* contribution of iron bound to neuromelanin of pigmented neurons, reinforcing the importance of imaging the substantia nigra in subregions with multi-contrast techniques. From our data, segmentation of R_2_*-based substantia nigra showed a similar ROI definition to the QSM (Supplementary Information Figure 2, Dice=0.7). We found that the Overlap ROI shares a similar location to the N1-sign obtained with SWI (Dice=0.3), but not completely (red and green outlines in Figure 3). Segmentation was performed on an independent healthy control group using the same imaging strategy as in the group study in MNI space with the purpose of reducing ROI selection bias.

Magnetization transfer contrast was reduced in Parkinson’s disease, suggestive of lower neuromelanin content as described in imaging and *post-mortem* studies (13,17,53,57–59). In addition, increased susceptibility-χ and R_2_* was found in patients in sub-regions driven by the MT-ROI, but not on regions exclusively showing high-susceptibility contrast in healthy controls. In our analyses, reduction of MT-CNR and increased QSM and R_2_* was most significant in the Overlap ROI. These results are supported by the ROC analysis (Figure 6). Our results also show that the N1-sign region alone does not show improved detectability of the disease in any of the imaging metrics studied. This indicated the importance of combining QSM and MT-w imaging for Parkinson’s disease diagnosis. Similar studies have reported the increase of susceptibility MR signal in the *pars compacta* region in patients with Parkinson’s disease (60–62) and, more recently, Bergsland et al. (63) narrowed it to the ventral portion. In (55) the hyperintensity loss in SWI images was better detected in the intermediate layer of the substantia nigra rich in dopaminergic neurons, the ventral portion of the SNpc, which in accordance with our definition of the Overlap ROI.

Distinguishing types of iron in the brain with MRI *in vivo* is challenging. Here we referred to R_2_*-to-χ ratio as an indirect estimate of the ferric (Fe^3+^) to ferrous (Fe^2+^) load in the brain. Our results show that, in patients, there is a significant increase of this ratio within the ventral portion of the SNpc. We differentiated iron markers in MRI in Parkinson’s patients by combining both magnitude and phase data from a single MRI acquisition. The R_2_*-to-χ ratio as a proxy for ferric to ferrous loading has been previously validated in phantom work using MRI in (25). However, in future, *post-mortem* MRI imaging together with histological iron staining is still required to validate this measure in healthy and diseased tissue.

Reports of the correlation of imaging and clinical outcomes in Parkinson’s disease are diverse. Some have shown disease duration has a positive correlation with susceptibility measures (QSM or R_2_*) in the substantia nigra (60,63–65) while others showed a weak to no correlation (22,48). When corrected for age, we found that only the R_2_*-to-χ ratio positively associated with disease duration, and most significantly in neuromelanin rich areas, including the Overlap ROI. Our results are in line with previous *post mortem* studies (Figure 1 from Youdim et al. (66)). Parkinson’s disease patients appear to show an imbalance of the Fe^3+^:Fe^2+^ ratio which is required for lipid peroxidation, resulting not only in an increased Fe^3+^-melanin complex in neuromelanin-pigmented neurons (67) but also of free iron because the neuromelanin buffering capacities towards iron became exhausted. This leads to toxicity and to oxygen radical-induced death of neuromelanin-pigmented dopaminergic cell populations. Histopathological studies such as (68,69) show a negative correlation between the number of pigmented neurons in the substantia nigra and clinical features such as disease duration or UPDRS-scores. In Biondetti et al. (70), neuromelanin-sensitive imaging correlated with motor, cognitive and mood/behavioural clinical scores in distinct regions of the substantia nigra. In our study, no neuromelanin markers correlated with clinical outcomes. This may be explained by the lack of granularity of our broad clinical measures compared to (70), and to the early stage of disease in our cohort (5±3 years from onset, Hoehn and Yahr 1.5-3) which may show a highly variable neuronal density loss within the substantia nigra: Kordower et al. (68) reported a wide range (33-80%) in the reduction of neuromelanin-containing neurons within 5-7 years post-PD diagnosis.

T_2_*-MRI (mathematical inverse of R_2_*) has been found to positively correlate with cognitive dysfunction in Parkinson’s disease in Tambasco et al. (71), but other studies have not found any strong correlations (72). From our data, patients that showed higher ferric loading (higher R_2_*-to-χ ratio) within the Overlap region definition of the substantia nigra had lower MoCA scores (i.e. worse cognition). Our data is consistent with the suggestion that the loss of neuromelanin-rich dopaminergic neurons and increased neurodegeneration lead to cognitive impairment. Neither R_2_* or QSM alone correlated with the MoCA score. This may be due to the lack of sensitivity of cognitive screening tests in non-demented patients and those without mild cognitive impairment, and also the specificity of the molecular correlates of our sequences in contrast to volumetric, spectroscopic or diffusion properties of other MR sequences. To associate our high-resolution MRI metrics with clinical outcomes, advanced stage patients may be required (65).

We demonstrate the usefulness of a neuromelanin-iron multimodal imaging approach. However, the addition of diffusion scalars similar to (73) at high-resolution would provide additional quantitative values of the microstructural integrity (e.g. myelin) and intra-to extra-cellular water diffusion variations within the nigra compartments that could aid characterization of the neurodegeneration in Parkinsonian disorders.

Hemispheric effects were found on both control and patient data, particularly evident on smaller ROIs (e.g. results for the SWIROI in Figure 5). Yet, we found no associations between imaging and clinical motor symptom laterality. The hemispheric asymmetry on the imaging results might be related to the known asymmetry in the B_1_^+^ transmit profile which occurs when using transmit-receive coils such as the coil used in our study in circular-polarised mode (74,75). In our analysis, we controlled for hemisphere effects by either adding a fixed variable of no interest on the general linear modelling statistics or by averaging the data across both hemispheres.

Magnetization transfer imaging shows specificity to the dopaminergic neurons of the SNpc but its biological relationship with neuromelanin is still elusive (76–78) as a contrast-to-noise measurement is only semi-quantitative. In order to better quantify nonexchangeable protons attached to macromolecules like neuromelanin, kinetic model parameters describing magnetization dynamics via a two-pool model could be used (79,80). Susceptibility and R2* mapping have the advantage of quantifying iron-related microscopic field changes, and so provide a more biologically meaningful clinical indicator of the pathogenesis. Therefore, the combination of these two imaging techniques provide a more holistic understanding of the neurodegeneration in the dopaminergic pathway within the substantia nigra and a more sensitive approach in differentiating patients with Parkinson’s disease from healthy controls.

In conclusion, the complementary information from iron and neuromelanin high-resolution ultra-high field 7T imaging allowed the detection of ferric (Fe^3+^) overload and neuromelanin loss on Parkinson’s patients in an area of the substantia nigra with both high neuromelanin and susceptibility MRI signal in normal controls. In addition, patients at later stages of the disease show higher ferric loading and lower cognitive function which is in accordance with histopathological findings. Our results suggest that ultra-high field 7T MT-weighted and T2*-weighted imaging are sensitive to the detection of molecular change in Parkinson’s disease, and may therefore support future experimental medicines strategies.

## Data Availability

The data and analysis scripts are data has been archived and is available from the authors upon request.

## Acknowledgements

We acknowledge the NIHR Cambridge Clinical Research Facility and the NIHR Cambridge Biomedical Research Centre (BRC-1215-20014), the Medical Research Council (MR/M008983/1, SUAG/051 G101400, MR/P01271X/1), the Wellcome Trust (103838), the Wellcome Trust & Royal Society (098436/Z/12/B), and the Cambridge Centre for Parkinson-plus (RG95450), Parkinson’s UK (K-1702), a Neil Hamilton Fairley Fellowship from the Australian National Health and Medical Research Council (GNT1091310), the Chinese Scholarship Council, a Cambridge Trust Vice-Chancellor’s Award and Fitzwilliam College. The views expressed are those of the authors and not necessarily those of the NHS, the NIHR or the Department of Health and Social Care. This is an open access article distributed under the Creative Commons Attribution License 4.0 (CCBY), which permits unrestricted use, distribution, and reproduction in any medium, provided the original work is properly cited.

## Authors Roles

Catarina Rua: Research Project (Conception, Organization, Execution), Statistical Analysis (Design, Execution, Review), Manuscript (Writing first draft, Review and Critique).

Claire O’Callaghan: Research Project (Conception, Organization, Execution), Statistical Analysis (Design, Review), Manuscript (Review and Critique).

Rong Ye: Research Project (Organization, Execution), Statistical Analysis (Design, Review), Manuscript (Review and Critique).

Frank H Hezemans: Research Project (Organization, Execution), Statistical Analysis (Review), Manuscript (Review and Critique).

Luca Passamonti: Research Project (Conception), Manuscript (Review and Critique).

P Simon Jones: Statistical Analysis (Design, Review), Manuscript (Review and Critique).

Guy B Williams: Research Project (Conception, Organization), Manuscript (Review and Critique).

Christopher T Rodgers: Research Project (Conception), Manuscript (Review and Critique). James B Rowe: Research Project (Conception, Organization), Statistical Analysis (Review), Manuscript (Review and Critique).

## Notes

### Competing Interest Statement

The authors have declared no competing interest.

### Author Declarations

The study was approved by the Cambridge Research Ethics Committee (16/EE/0084; 10/H0308/34) and all participants provided written informed consent in accordance with the Declaration of Helsinki.

## References

1. Hirsch EC, Brandel J -P, Galle P, Javoy-Agid F, Agid Y. Iron and Aluminum Increase in the Substantia Nigra of Patients with Parkinson’s Disease: An X-Ray Microanalysis. J Neurochem. 1991;56(2):446–51.

2. Kitao S, Matsusue E, Fujii S, Miyoshi F, Kaminou T, Kato S, et al. Correlation between pathology and neuromelanin MR imaging in Parkinson’s disease and dementia with Lewy bodies. Neuroradiology. 2013;55(8):947–53.

3. Vaillancourt DE, Mitchell T. Parkinson’s disease progression in the substantia nigra: location, location, location. Brain. 2020;143(9):2728–2630.

4. Zecca L, Wilms H, Geick S, Claasen JH, Brandenburg LO, Holzknecht C, et al. Human neuromelanin induces neuroinflammation and neurodegeneration in the rat substantia nigra: Implications for Parkinson’s disease. Acta Neuropathol. 2008;116(1):47–55.

5. Götz ME, Double K, Gerlach M, Youdim MBH, Riederer P. The relevance of iron in the pathogenesis of Parkinson’s disease. Ann N Y Acad Sci. 2004;1012(March):193–208.

6. Double KL, Ben-Shachar D, Youdim MBH, Zecca L, Riederer P, Gerlach M. Influence of neuromelanin on oxidative pathways within the human substantia nigra. Neurotoxicol Teratol. 2002;24(5):621–8.

7. Eid R, Arab NTT, Greenwood MT. Iron mediated toxicity and programmed cell death: A review and a re-examination of existing paradigms. Biochim Biophys Acta - Mol Cell Res [Internet]. 2017;1864(2):399–430. Available from: http://dx.doi.org/10.1016/j.bbamcr.2016.12.002

8. Youdim MB. Iron in the brain: implications for Parkinson’s and Alzheimer’s diseases. Mt Sinai J Med [Internet]. 1988 Jan;55(1):97—101. Available from: http://europepmc.org/abstract/MED/3279308

9. Braak H, Del Tredici K, Rüb U, De Vos RAI, Jansen Steur ENH, Braak E. Staging of brain pathology related to sporadic Parkinson’s disease. Neurobiol Aging. 2003;24(2):197–211.

10. Zucca FA, Segura-Aguilar J, Ferrari E, Muñoz P, Paris I, Sulzer D, et al. Interactions of iron, dopamine and neuromelanin pathways in brain aging and Parkinson’s disease. Prog Neurobiol. 2017;155:96–119.

11. Zucca FA, Bellei C, Giannelli S, Terreni MR, Gallorini M, Rizzio E, et al. Neuromelanin and iron in human locus coeruleus and substantia nigra during aging: Consequences for neuronal vulnerability. J Neural Transm. 2006;113(6):757–67.

12. Grifiths PD, Dobson BR, Jones GR, Clarke DT. Iron in the basal ganglia in Parkinson’s disease. An in vivo study using extended X-ray absorption fine structure and cryo-electron microscopy. Brain. 1999;122(4):667–73.

13. Isaias IU, Trujillo P, Summers P, Marotta G, Mainardi L, Pezzoli G, et al. Neuromelanin imaging and dopaminergic loss in parkinson’s disease. Front Aging Neurosci. 2016;8(AUG):1–12.

14. Wakamatsu K, Fujikawa K, Zucca FA, Zecca L, Ito S. The structure of neuromelanin as studied by chemical degradative methods. J Neurochem. 2003;86(4):1015–23.

15. Sasaki M, Shibata E, Tohyama K, Takahashi J, Otsuka K, Tsuchiya K, et al. Neuromelanin magnetic resonance imaging of locus ceruleus and substantia nigra in Parkinson’s disease. Neuroreport. 2006;17(11):1215–8.

16. Reimão S, Ferreira S, Nunes RG, Pita Lobo P, Neutel D, Abreu D, et al. Magnetic resonance correlation of iron content with neuromelanin in the substantia nigra of early-stage Parkinson’s disease. Eur J Neurol. 2016;23(2):368–74.

17. Prasad S, Stezin A, Lenka A, George L, Saini J, Yadav R, et al. Three-dimensional neuromelanin-sensitive magnetic resonance imaging of the substantia nigra in Parkinson’s disease. Eur J Neurol. 2018;25(4):680–6.

18. Keren NI, Taheri S, Vazey EM, Morgan PS, Granholm ACE, Aston-Jones GS, et al. Histologic validation of locus coeruleus MRI contrast in post-mortem tissue. Neuroimage [Internet]. 2015;113:235–45. Available from: http://dx.doi.org/10.1016/j.neuroimage.2015.03.020

19. Haacke EM, Cheng NYC, House MJ, Liu Q, Neelavalli J, Ogg RJ, et al. Imaging iron stores in the brain using magnetic resonance imaging. Magn Reson Imaging. 2005;23(1):1–25.

20. Lotfipour AK, Wharton S, Schwarz ST, Gontu V, Schäfer A, Peters AM, et al. High resolution magnetic susceptibility mapping of the substantia nigra in Parkinson’s disease. J Magn Reson Imaging. 2012;35(1):48–55.

21. Barbosa JHO, Santos AC, Tumas V, Liu M, Zheng W, Haacke EM, et al. Quantifying brain iron deposition in patients with Parkinson’s disease using quantitative susceptibility mapping, R2 and R2*. Magn Reson Imaging [Internet]. 2015;33(5):559–65. Available from: http://dx.doi.org/10.1016/j.mri.2015.02.021

22. Acosta-Cabronero J, Cardenas-Blanco A, Betts MJ, Butryn M, Valdes-Herrera JP, Galazky I, et al. The whole-brain pattern of magnetic susceptibility perturbations in Parkinson’s disease. Brain. 2017;140(1):118–31.

23. Rua C, Clarke WT, Driver ID, Mougin O, Morgan AT, Clare S, et al. Multi-centre, multi-vendor reproducibility of 7T QSM and R2* in the human brain: Results from the UK7T study. Neuroimage [Internet]. 2020;223(April):117358. Available from: https://doi.org/10.1016/j.neuroimage.2020.117358

24. Cheng Z, He N, Huang P, Li Y, Tang R, Sethi SK, et al. Imaging the Nigrosome 1 in the substantia nigra using susceptibility weighted imaging and quantitative susceptibility mapping: An application to Parkinson’s disease. NeuroImage Clin [Internet]. 2020;25(May 2019):102103. Available from: https://doi.org/10.1016/j.nicl.2019.102103

25. Dietrich O, Levin J, Giese A, Plate A, Bötzel K, Reiser MF, et al. Differentiation of Fe2+ and Fe3+ with iron-sensitive MRI. In: Proceedings 22nd Scientific Meeting, International Society for Magnetic Resonance in Medicine. 2013. p. 2482.

26. Priovoulos N, Jacobs HIL, Ivanov D, Uludağ K, Verhey FRJ, Poser BA. High-resolution in vivo imaging of human locus coeruleus by magnetization transfer MRI at 3T and 7T. Neuroimage [Internet]. 2018;168(July):427–36. Available from: http://dx.doi.org/10.1016/j.neuroimage.2017.07.045

27. Ye R, Rua C, O’Callaghan C, Jones PS, Hezemans FH, Kaalund SS, et al. An in vivo probabilistic atlas of the human locus coeruleus at ultra-high field. Neuroimage [Internet]. 2021;225(September 2020):117487. Available from: https://doi.org/10.1016/j.neuroimage.2020.117487

28. Tokuhiro T, Appleby A, Leghrouz A, Metcalf R, Tokarz R. Proton spin-lattice relaxation of water molecules in ferrous-ferric/agarose gel system. J Chem Phys. 1996;105(9):3761–9.

29. Martinez-Martin P, Falup-Pecurariu C, Rodriguez-Blazquez C, Serrano-Dueñas M, Carod Artal FJ, Rojo Abuin JM, et al. Dementia associated with Parkinson’s disease: Applying the Movement Disorder Society Task Force criteria. Park Relat Disord [Internet]. 2011;17(8):621–4. Available from: http://dx.doi.org/10.1016/j.parkreldis.2011.05.017

30. Weintaub D, Koester J, Potenza MN, Siderowf AD, Stacy M, Voon V, et al. Impulse Control Disorders in Parkinson Disease. Arch Neurol. 2010;67(5):589–95.

31. Tomlinson CL, Stowe R, Patel S, Rick C, Gray R, Clarke CE. Systematic review of levodopa dose equivalency reporting in Parkinson’s disease. Mov Disord. 2010;25(15):2649–53.

32. Goetz CG, Nutt JG, Stebbins GT. The unified dyskinesia rating scale: Presentation and clinimetric profile. Mov Disord. 2008;23(16):2398–403.

33. Folstein MF, Folstein SE. Mini-Mental State: a practical method for grading the cognitive state of patients for the clinician. Int J Geriatr Psychiatry. 1975;12:189–98.

34. Nasreddine ZS, Phillips NA, Bédirian V, Charbonneau S, Whitehead V, Collin I, et al. The Montreal Cognitive Assessment, MoCA: A brief screening tool for mild cognitive impairment. J Am Geriatr Soc. 2005;53(4):695–9.

35. Zigmond AS, Snaith RP. The Hospital Anxiety and Depression Scale. Acta Psychiatr Scand. 1983;67:361–70.

36. Patton JH, Stanford MS, Barratt ES. Factor structure of the barratt impulsiveness scale. J Clin Psychol. 1995;51(6):768–74.

37. Starkstein SE, Mayberg HS, Preziosi TJ, Andrezejewski P, Leiguarda R, Robinson RG. Reliability, validity, and clinical correlates of apathy in Parkinson’s disease. J Neuropsychiatry Clin Neurosci. 1992;4(2):134–9.

38. Roemer PB, Edelstein WA, Hayes CE, Souza SP, Mueller OM. The NMR phased array. Magn Reson Med. 1990;16(2):192–225.

39. Mougin O, Clarke WT, Driver I, Rua C, Morgan AT, Francis S, et al. Robustness of PSIR segmentation and R1 mapping at 7T: a travelling head study. In: Proceedings 27th Scientific Meeting, International Society for Magnetic Resonance in Medicine. Paris; 2017. p. 237.

40. Acosta-Cabronero J, Milovic C, Mattern H, Tejos C, Speck O, Callaghan MF. A robust multiscale approach to quantitative susceptibility mapping. Neuroimage [Internet]. 2018;183(June):7–24. Available from: https://doi.org/10.1016/j.neuroimage.2018.07.065

41. Pei M, Nguyen TD, Thimmappa ND, Salustri C, Dong F, Cooper MA, et al. Algorithm for fast monoexponential fitting based on Auto-Regression on Linear Operations (ARLO) of data. Magn Reson Med. 2015;73(2):843–50.

42. Li N, Wang W-T, Sati P, Pham DL, Butman JA. Quantitative assessment of susceptibility weighted imaging processing methods. J Magn Reson Imaging [Internet]. 2014;40(6):1463–73. Available from: https://www.ncbi.nlm.nih.gov/pmc/articles/PMC3624763/pdf/nihms412728.pdf

43. Clarke WT, Mougin O, Driver ID, Rua C, Morgan AT, Asghar M, et al. Multi-site harmonization of 7 tesla MRI neuroimaging protocols. Neuroimage [Internet]. 2020;206(October 2019):116335. Available from: https://doi.org/10.1016/j.neuroimage.2019.116335

44. Fonov V, Evans AC, Botteron K, Almli CR, McKinstry RC, Collins DL. Unbiased average age-appropriate atlases for pediatric studies. Neuroimage [Internet]. 2011;54(1):313–27. Available from: http://dx.doi.org/10.1016/j.neuroimage.2010.07.033

45. Avants BB, Epstein CL, Grossman M, Gee JC. Symmetric diffeomorphic image registration with cross-correlation: Evaluating automated labeling of elderly and neurodegenerative brain. Med Image Anal. 2008;12(1):26–41.

46. Langley J, Huddleston DE, Sedlacik J, Boelmans K, Hu XP. Parkinson’s disease–related increase of T2*-weighted hypointensity in substantia nigra pars compacta. Mov Disord. 2017;32(3):441–9.

47. Chen X, Huddleston DE, Langley J, Ahn S, Barnum CJ, Factor SA, et al. Simultaneous imaging of locus coeruleus and substantia nigra with a quantitative neuromelanin MRI approach. Magn Reson Imaging [Internet]. 2014;32(10):1301–6. Available from: http://dx.doi.org/10.1016/j.mri.2014.07.003

48. He N, Ghassaban K, Huang P, Jokar M, Wang Y, Cheng Z, et al. Imaging Iron and Neuromelanin Simultaneously Using a Single 3D Magnetization Transfer Sequence: Combining Neuromelanin, Iron and the Nigrosome-1 Sign as Complementary Imaging Biomarkers in Early Stage Parkinson’s Disease. Neuroimage [Internet]. 2021;21:117810. Available from: https://doi.org/10.1016/j.neuroimage.2021.117810

49. Yushkevich PA, Piven J, Hazlett HC, Smith RG, Ho S, Gee JC, et al. User-guided 3D active contour segmentation of anatomical structures: Significantly improved efficiency and reliability. Neuroimage. 2006;31(3):1116–28.

50. Eugenio J, Leemput K Van, Bhatt P, Casillas C, Dutt S, Schuff N, et al. NeuroImage Bayesian segmentation of brainstem structures in MRI. Neuroimage [Internet]. 2015;113:184–95. Available from: http://dx.doi.org/10.1016/j.neuroimage.2015.02.065

51. Singmann H, Kellen D. An Introduction to Mixed Models for Experimental Psychology. New Methods Cogn Psychol. 2019;4–31.

52. Langley J, Huddleston DE, Chen X, Sedlacik J, Zachariah N, Hu X. A multicontrast approach for comprehensive imaging of substantia nigra. Neuroimage. 2015;112(404):7–13.

53. Damier P, Hirsch EC, Agid Y, Graybiel AM. The substantia nigra of the human brain: II. Patterns of loss of dopamine-containing neurons in Parkinson’s disease. Brain. 1999;122(8):1437–48.

54. Nieuwenhuys R, Voogd D, van Huijzen C. The Human Central Nervous System. Springer-Verlag. Germany; 2008.

55. Cosottini M, Frosini D, Pesaresi I, Costagli M, Biagi L, Cerabolo R, et al. MR Imaging of the Substantia Nigra at 7 T Enables Diagnosis. Radiology. 2014;271(3):831–8.

56. Brammerloh M, Morawski M, Weigelt I, Reinert T, Lange C, Pelicon P, et al. Measuring the iron content of dopaminergic neurons in substantia nigra with MRI relaxometry. bioRxiv. 2020;1:1–25.

57. Fearnley JM, Lees AJ. Ageing and Parkinson’s disease: substantia nigra regional selectivity. Brain. 1991;114(1):2283–301.

58. Takahashi H, Watanabe Y, Tanaka H, Mihara M, Mochizuki H, Takahashi K, et al. Comprehensive MRI quantification of the substantia nigra pars compacta in Parkinson’s disease. Eur J Radiol [Internet]. 2018;109(May):48–56. Available from: https://doi.org/10.1016/j.ejrad.2018.06.024

59. Ohtsuka C, Sasaki M, Konno K, Kato K, Takahashi J, Yamashita F, et al. Differentiation of early-stage parkinsonisms using neuromelanin-sensitive magnetic resonance imaging. Park Relat Disord [Internet]. 2014;20(7):755–60. Available from: http://dx.doi.org/10.1016/j.parkreldis.2014.04.005

60. Du G, Liu T, Lewis MM, Kong L, Wang Y, Connor J, et al. Quantitative susceptibility mapping of the midbrain in Parkinson’s disease. Mov Disord. 2016;31(3):317–24.

61. Chen Q, Chen Y, Zhang Y, Wang F, Yu H, Zhang C, et al. Iron deposition in Parkinson’s disease by quantitative susceptibility mapping. BMC Neurosci [Internet]. 2019;20(1):1–8. Available from: https://doi.org/10.1186/s12868-019-0505-9

62. Lotfipour AK, Wharton S, Schwarz ST, Gontu V, Schäfer A, Peters AM, et al. High resolution magnetic susceptibility mapping of the substantia nigra in Parkinson’s disease. J Magn Reson Imaging. 2012;35(1):48–55.

63. Bergsland N, Zivadinov R, Schweser F, Hagemeier J, Lichter D, Guttuso T. Ventral posterior substantia nigra iron increases over 3 years in Parkinson’s disease. Mov Disord. 2019;34(7):1006–13.

64. He N, Ling H, Ding B, Huang J, Zhang Y, Zhang Z, et al. Region-specific disturbed iron distribution in early idiopathic Parkinson’s disease measured by quantitative susceptibility mapping. Hum Brain Mapp. 2015;36(11):4407–20.

65. Guan X, Xuan M, Gu Q, Huang P, Liu C, Wang N, et al. Regionally progressive accumulation of iron in Parkinson’s disease as measured by quantitative susceptibility mapping. NMR Biomed. 2017;30(4).

66. Youdim MBH, Ben-Shachar D, Riederer P. Is Parkinson’s disease a progressive siderosis of substantia nigra resulting in iron and melanin induced neurodegeneration? Acta Neurol Scand. 1989;80(1):47–54.

67. Jellinger K, Kienzl E, Rumpelmair G, Riederer P, Stachelberger H, Ben-Shachar D, et al. Iron-Melanin Complex in Substantia Nigra of Parkinsonian Brains: An X-Ray Microanalysis. J Neurochem. 1992;59(3):1168–71.

68. Kordower JH, Olanow CW, Dodiya HB, Chu Y, Beach TG, Adler CH, et al. Disease duration and the integrity of the nigrostriatal system in Parkinson’s disease. Brain. 2013;136(8):2419–31.

69. Greffard S, Verny M, Bonnet AM, Beinis JY, Gallinari C, Meaume S, et al. Motor score of the unified Parkinson disease rating scale as a good predictor of lewy body-associated neuronal loss in the substantia nigra. Arch Neurol. 2006;63(4):584–8.

70. Biondetti E, Gaurav R, Yahia-Cherif L, Mangone G, Pyatigorskaya N, Valabrègue R, et al. Spatiotemporal changes in substantia nigra neuromelanin content in Parkinson’s disease. Brain. 2020;143(9):2757–70.

71. Tambasco N, Paolini Paoletti F, Chiappiniello A, Lisetti V, Nigro P, Eusebi P, et al. T2*-weighted MRI values correlate with motor and cognitive dysfunction in Parkinson’s disease. Neurobiol Aging [Internet]. 2019;80:91–8. Available from: https://doi.org/10.1016/j.neurobiolaging.2019.04.005

72. Zhang W, Sun SG, Jiang YH, Qiao X, Sun X, Wu Y. Determination of brain iron content in patients with Parkinson’s disease using magnetic susceptibility imaging. Neurosci Bull. 2009;25(6):353–60.

73. Péran P, Cherubini A, Assogna F, Piras F, Quattrocchi C, Peppe A, et al. Magnetic resonance imaging markers of Parkinson’s disease nigrostriatal signature. Brain. 2010;133(11):3423–33.

74. Vaidya M V, Collins CM, Sodickson DK, Brown R, Wiggins GC, Lattanzi R. Dependence of B1+ and B1-Field Patterns of Surface Coils on the Electrical Properties of the Sample and the MR Operating Frequency. Concepts Magn Reson Part B Magn Reson Eng. 2016;46B(1):25–40.

75. Malik SJ, Beqiri A, Padormo F, Hajnal J V. Direct Signal Control of the steady-state response of 3D-FSE sequences. Magn Reson Med. 2015;73(3):951–63.

76. Cassidy CM, Zucca FA, Girgis RR, Baker SC, Weinstein JJ, Sharp ME, et al. Neuromelanin-sensitive MRI as a noninvasive proxy measure of dopamine function in the human brain. Proc Natl Acad Sci U S A. 2019;116(11):5108–17.

77. Priovoulos N, van Boxel SCJ, Jacobs HIL, Poser BA, Uludag K, Verhey FRJ, et al. Unraveling the contributions to the neuromelanin-MRI contrast. Brain Struct Funct [Internet]. 2020;225(9):2757–74. Available from: https://doi.org/10.1007/s00429-020-02153-z

78. Watanabe T, Tan Z, Wang X, Martinez-Hernandez A, Frahm J. Magnetic resonance imaging of noradrenergic neurons. Brain Struct Funct [Internet]. 2019;224(4):1609–25. Available from: http://dx.doi.org/10.1007/s00429-019-01858-0

79. Henkelman RM, Stanisz GJ, Graham SJ. Magnetization transfer in MRI: A review. NMR Biomed. 2001;14(2):57–64.

80. Yarnykh VL. Fast macromolecular proton fraction mapping from a single off-resonance magnetization transfer measurement. Magn Reson Med. 2012;68(1):166–78.

