## Supplementary Information for "Substantia nigra ferric overload and neuromelanin loss in Parkinson’s disease measured with 7T MRI"

C Rua et al.

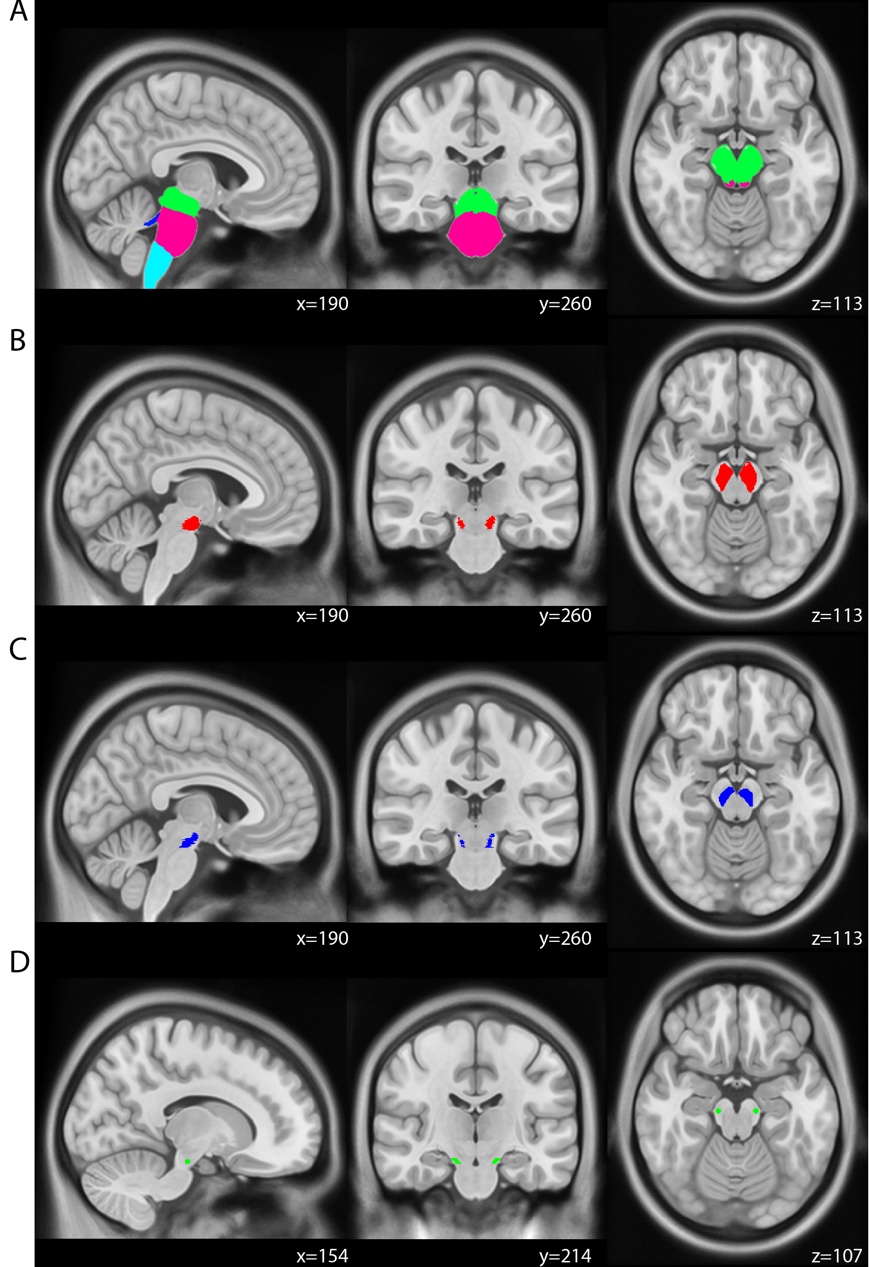

**Supplementary Information Figure 1: (A) Brainstem segmentation ROIs obtained in Freesurfer 6.0: midbrain (green), pons (pink), medulla (light blue) and superior cerebellar peduncle (dark blue). (B) Left and right search areas for the QSM SN ROI definition. (C) Left and right search areas for the MT SN ROI definition. (D) Left and right cylindrical reference ROIs in the midbrain.**

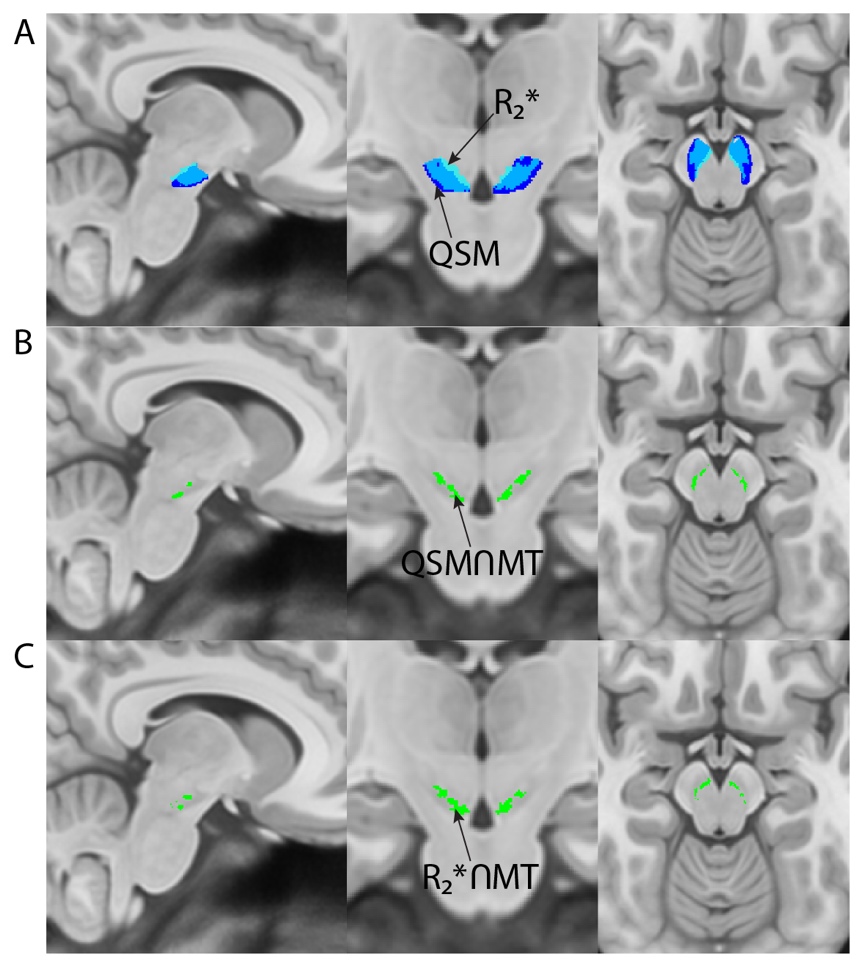

**Supplementary Information Figure 2: Sagittal, coronal and axial views of MNI template showing ROI definitions based on QSM and R_2_* data. (A) SN region of interest based on QSM (dark blue) and R_2_* (light blue) data. R_2_* ROI is overlayed on QSM ROI with 30% transparency to observe the overlapping region (Dice-index=0.7). (B) ‘Overlap’ ROI (green) based on the QSMROI and MTROI. (C) ‘Overlap’ ROI (green) based on the R_2_* ROI and MTROI. (B) and (C) show similar definitions of the overlapping region with the MTROI.**

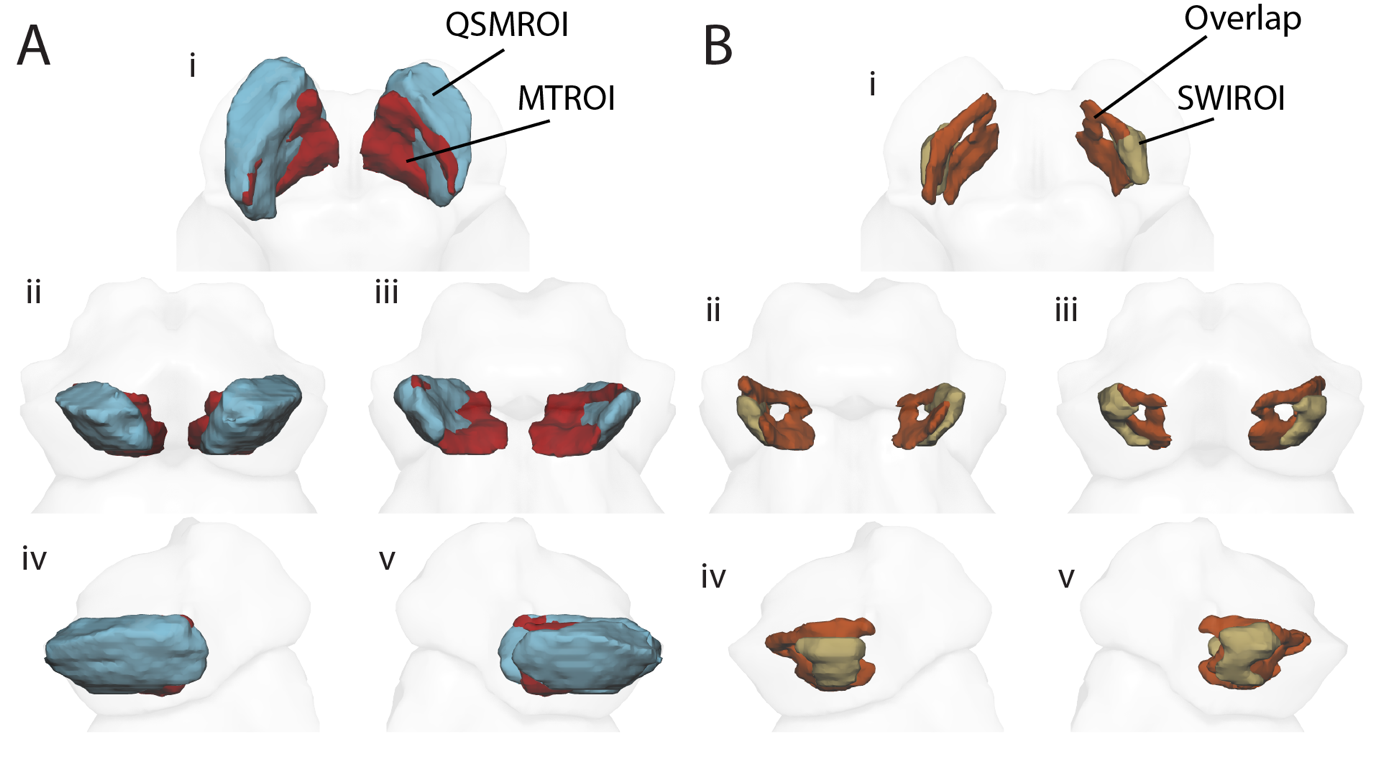

**Supplementary Information Figure 3: 3D rendering of the MTROI (blue) and QSMROI (red) (A), and Overlap (orange) and SWIROI (brown) (B) ROIs in the substantia nigra in the midbrain in ICBM 152 template space. Views: i: superior; ii: anterior; iii: posterior; iv: left; v: right.**

| **Imaging Metric** | **ROI** | **Average±Standard Deviation*** | **p-value (HC vs. PD)** |
| --- | --- | --- | --- |
| MT-CNR | MTROI | HC: 4.9±2.0  PD: 2.8±1.5 | **< .0001** |
|  | MTOnly | HC: 5.1±2.0  PD: 3.4±1.5 | **< .0001** |
|  | Overlap | HC: 4.6±2.1  PD: 2.4±1.8 | **< .0001** |
|  | QSMROI | HC: 1.6±1.4  PD: 0.4±1.3 | **.0013** |
|  | QSMOnly | HC: 0.9±1.3  PD: 0.003±1.3 | **.012** |
|  | SWIROI | HC: 2.9±1.4  PD: 1.6±1.1 | **< .0001** |

**Supplementary Information Table 1: Summary statistics and p-values of group comparisons for the MT-CNR in each of the six SN ROIs. *The left and right hemisphere data was averaged.**

| **Imaging Metric** | **ROI** | **Average±Standard Deviation*** | **p-value (HC vs. PD)** |
| --- | --- | --- | --- |
| QSM $\chi$  (ppm) | MTROI | HC: 0.089±0.029  PD: 0.120±0.018 | **< .0001** |
|  | MTOnly | HC: 0.054±0.034  PD: 0.079±0.027 | **< .0001** |
|  | Overlap | HC: 0.12±0.032  PD: 0.15±0.029 | **< .0001** |
|  | QSMROI | HC: 0.097±0.029  PD: 0.097±0.026 | .96 |
|  | QSMOnly | HC: 0.092±0.031  PD: 0.084±0.028 | 0.29 |
|  | SWIROI | HC: 0.089±0.029  PD: 0.098±0.031 | 0.27 |
| R_2_*  (ms) | MTROI | HC: 0.054±0.009  PD: 0.063±0.007 | **< .0001** |
|  | MTOnly | HC: 0.048±0.010  PD: 0.053±0.007 | **.0061** |
|  | Overlap | HC: 0.059±0.011  PD: 0.070±0.010 | **< .0001** |
|  | QSMROI | HC: 0.062±0.009  PD: 0.064±0.008 | .34 |
|  | QSMOnly | HC: 0.062±0.009  PD: 0.062±0.008 | .93 |
|  | SWIROI | HC: 0.052±0.010  PD: 0.058±0.008 | **.01** |
| $\Delta R_{2}^{*}/\Delta\chi$  (ms/ppm) | MTROI | HC: 0.21±0.078  PD: 0.23±0.052 | .13 |
|  | MTOnly | HC: 0.23±0.64  PD: 0.24±0.082 | .87 |
|  | Overlap | HC: 0.19±0.058  PD: 0.22±0.059 | **.0048** |
|  | QSMROI | HC: 0.27±0.071  PD: 0.30±0.090 | .058 |
|  | QSMOnly | HC: 0.30±0.084  PD: 0.34±0.13 | .054 |
|  | SWIROI | HC: 0.40±0.26  PD: 0.45±0.27 | .21 |

**Supplementary Information Table 2: Summary statistics and p-values of group comparisons for the T_2_*-w based metrics in each of the 5 SN ROIs. *The left and right hemisphere data was averaged.**

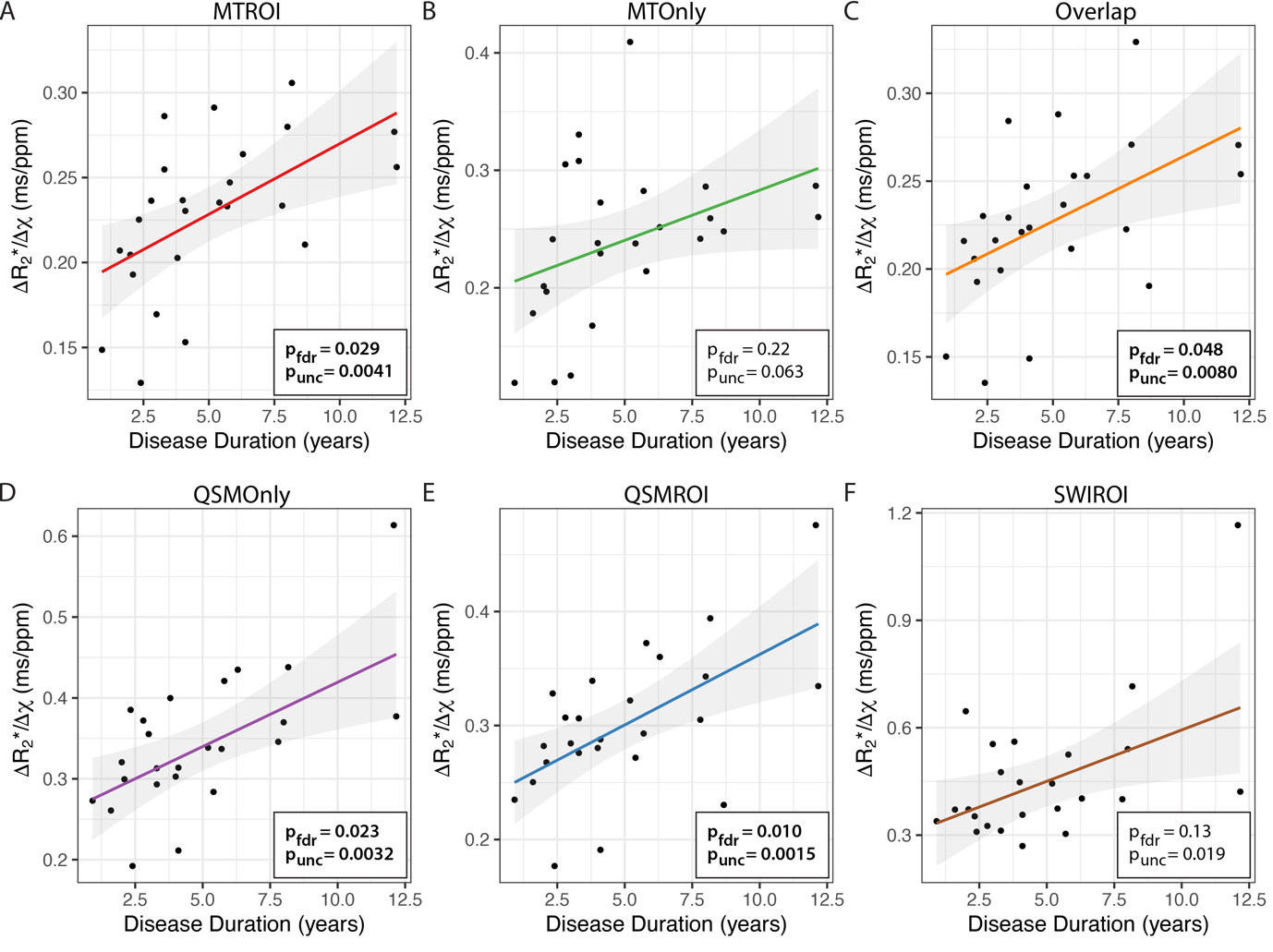

**Supplementary Information Figure 4: Associations of** $\boldsymbol{\Delta}\boldsymbol{R}_{\mathbf{2}}^{\mathbf{*}}\boldsymbol{/\Delta}\boldsymbol{\chi}$**ratio with disease duration in years in the Parkinson’s disease group, for each of the six sub-regions of the substantia nigra. Uncorrected and FDR-corrected p-values are reported in each plot for the linear fits.**

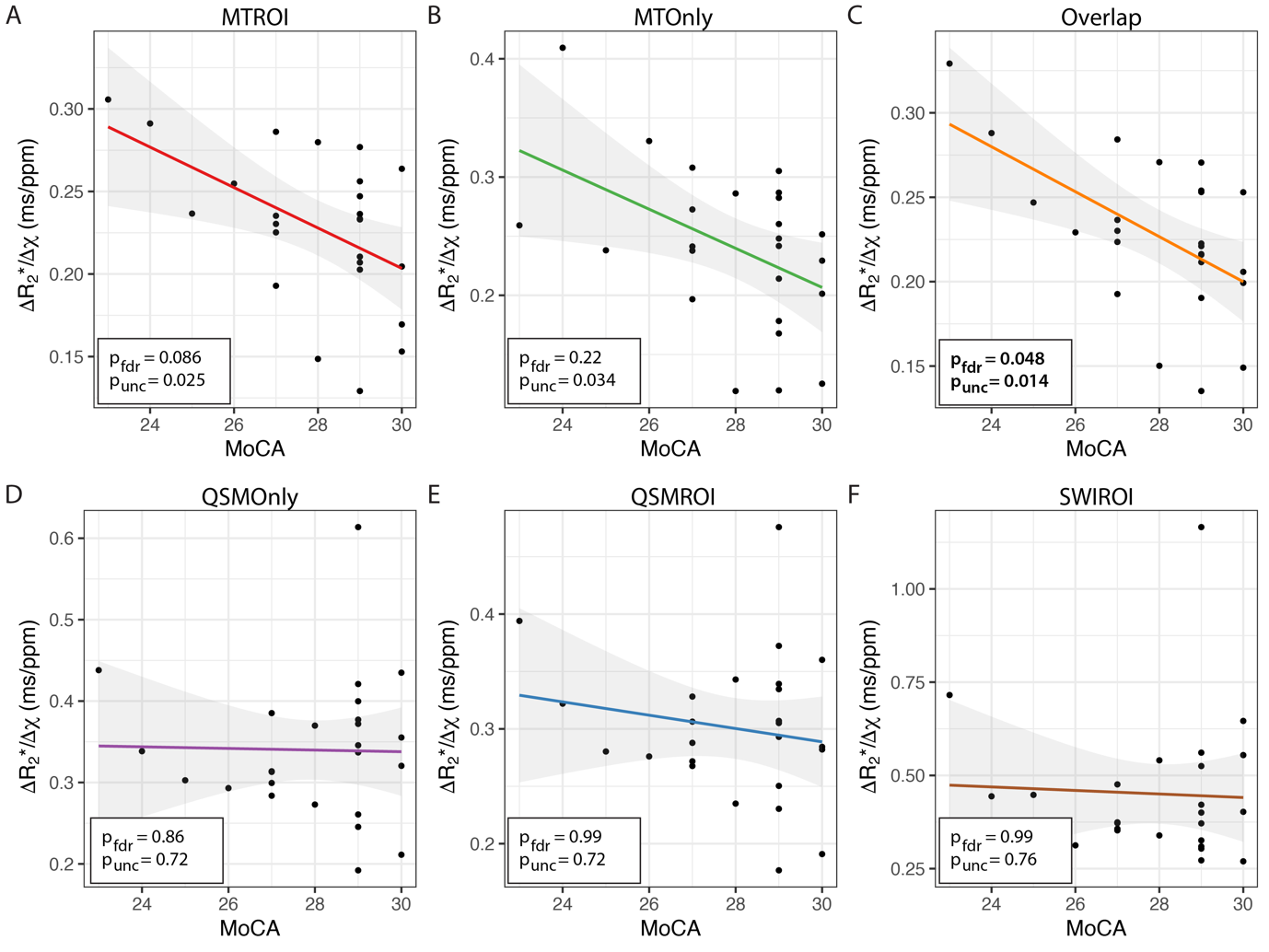

**Supplementary Information Figure 5: Associations of** $\boldsymbol{\Delta}\boldsymbol{R}_{\mathbf{2}}^{\mathbf{*}}\boldsymbol{/\Delta}\boldsymbol{\chi}$**ratio with the MoCA score in the Parkinson’s disease group, for each of the six sub-regions of the substantia nigra. Uncorrected and FDR-corrected p-values are reported in each plot for the linear fits.**

**Supplementary Information: Motor symptom and imaging laterality indexes**

Using the items from the MDS-UPDRS-III that have separate scores for the left and right side of the body, we calculated a motor asymmetry index (MAI). For each patient, MAI was calculated as:

$$\frac{\text{Right side symptoms}-\text{Left side symptoms}}{\text{Right side symptoms + Left side symptoms}}$$

Results from a one-sample t-test showed that the distribution of people with left vs. right dominant motor symptoms did not differ from zero (M = -0.01, SD = 0.40; *t*_(24)_ = -0.12, p = .904) resulting in no motor symptom laterality bias in our cohort.

For each patient and in each subregion of the substantia nigra, imaging asymmetry indexes (AI) were calculated similarly to MAI for MT-CNR, $\Delta\chi$, R_2_*, $\Delta R_{2}^{*}/\Delta\chi$ ratio using right and left hemisphere data, e.g.:

$$\Delta\chi AI=\frac{{\Delta\chi}_{right hemisphere}-{\Delta\chi}_{left hemisphere}}{{\Delta\chi}_{right hemisphere}+{\Delta\chi}_{left hemisphere}}$$
